# DHDDS-related juvenile parkinsonism is caused by impaired lipid metabolism, glycosylation, and mitochondrial dysfunction, which can be rescued by NAD⁺ treatment

**DOI:** 10.64898/2026.05.28.26354198

**Authors:** I.J.J. Muffels, K.A. Kantautas, G. MacDonald, K. Garapati, R.R. Pasupuleti, R.J. Tinker, R. Shah, M.A. Thevandavakkam, J. Donnelly, R. Hrstka, D. Smith, J. Van Klinken, F. Vaz, A. Pandey, E.O. Perlstein, T. Kozicz, E. Morava

## Abstract

**Background:** Mono-allelic Dehydrodolichyl Diphosphate Synthase (*DHDDS)* variants are associated with juvenile Parkinsonism, developmental delay and seizures. Symptoms are progressive, and various mechanisms, such as defective glycosylation, lysosomal dysfunction and cholesterol accumulation have been hypothesized to underlie disease symptoms. There is no treatment for DHDDS-related disease.

**Methods:** Patient-derived cortical forebrain organoids were created to elucidate disease mechanisms and evaluate potential treatments. In these neuronal models, glycosylation, lipidomics, proteomics, cholesterol/ganglioside accumulation, mitochondrial function and electrophysiological activity were assessed. Finally, we investigated the effects of nicotinamide mononucleotide (NMN), identified through a yeast-based drug screen, in neuronal cell models and in six patients in an off-label, N-of-1, observational series.

**Results:** DHDDS-patient derived organoids showed visual signs of degeneration after four months of culturing. This was accompanied by significant cholesterol accumulation in astrocytes, decreased mitochondrial respiration and loss of deep-layer neurons. In addition, we identified glycosylation abnormalities, showing for the first time that glycosylation in human tissue is affected by monoallelic *DHDDS* variants. Proteomic analysis revealed altered protein expression of proteins involved in lipid metabolism, cytoskeletal organization and neuronal development. We found that oral Nicotinamide Mononucleotide supplementation led to significant improvement in mitochondrial respiration and electrophysiological parameters in organoids, concurring with clinical improvements in all of the treated patients, particularly regarding their ataxia and tremor.

**Conclusion:** Our findings reveal a progressive phenotype in DHDDS-patient-derived brain organoids, with mitochondrial dysfunction and astrocyte-specific metabolic alterations contributing to disease pathology. Notably, NMN treatment led to clinical improvements in patients with heterozygous *DHDDS* variants, highlighting its potential as a therapeutic strategy.

## Introduction

Pathogenic heterozygous variants in Dehydrodolichyl Diphosphate Synthase (DHDDS) cause a progressive neurological disorder characterized by seizures, mixed movement abnormalities, and neurodevelopmental impairment.^1^ Disease onset and progression are highly variable, ranging from childhood to late adulthood, and the reason behind this heterogeneity remains largely unknown.^2^ Up to date, no disease-modifying therapies exist, and affected individuals often develop progressive disabilities. Clinically, DHDDS-associated disease overlaps with juvenile and early-onset parkinsonism syndromes, that show epilepsy, cognitive impairment, dystonia, and pyramidal signs, together with parkinsonism. As such, elucidating disease mechanisms in DHDDS-associated disease may provide broader insight into the pathobiology of juvenile parkinsonism. The progressive course of DHDDS-associated disease, unlike most glycosylation disorders, suggests that mechanisms beyond defective glycosylation, potentially involving disrupted lipid and lysosomal homeostasis, contribute to neurodegeneration.

DHDDS operates together with NgBR (encoded by the NUS1 gene) as subunits of the cis-prenyltransferase (*cis*-PTase) complex and utilizes isoprenoid intermediates from the mevalonate pathway, which also supplies isoprenoids for cholesterol biosynthesis.^3–6^ *DHDDS* encodes the catalytic subunit of *cis*-PTase responsible for synthesizing long-chain polyprenyl diphosphates that are converted to dolichol, an essential lipid carrier for protein glycosylation.^7,8^ Despite the essential role of DHDDS in glycosylation, glycosylation abnormalities have not been observed in patients with heterozygous variants in DHDDS^9^, although they have been reported in patients with the recessive form of the disease.^10^ Recently, Da Silva et al., found a significant decrease in oligomannose-containing N-glycans in mice carrying the heterozygous p.R37H *dhdds* variant, suggesting that glycosylation deficits might be present in organs affected by the disease, although this remains to be verified in humans.^11^

Beyond its role in protein glycosylation, additional mechanisms have been implicated in heterozygous DHDDS-associated disease. Unlike most glycosylation disorders, which stabilize or improve over time¹⁴, neurological manifestations in individuals with heterozygous *DHDDS* variants are typically progressive.^2^ This disparity suggests that glycosylation defects alone may not fully account for disease progression and that secondary metabolic disturbances may contribute to the phenotype. One proposed mechanism involves disrupted lipid homeostasis, including cholesterol and ganglioside accumulation, that has been observed in patient-derived skin fibroblasts with heterozygous *DHDDS* variants.^9^ In support of this, lysosomes containing intramembranous lipid whorls – indicative of glycosphingolipid storage – have been identified in nerve cells from patient-derived skin biopsies.^9,12^ However, cholesterol or glycosphingolipid accumulation has not been detected in the brains of 6-week-old heterozygous *dhdds* p.R37H mouse models.^11^ This discrepancy between mice and human skin biopsies highlights a key gap in our understanding of heterozygous DHDDD-associated disease. Are altered lipid homeostasis and lysosomal dysfunction key aspects of DHDDS-mediated disease mechanisms or are other components responsible for the pathophysiology?

Here, we studied Cortical Brain Organoids (CBOs) of patients with heterozygous *DHDDS* variants, and found that these variants drive a progressive, brain-specific disruption of lipid trafficking and lysosomal homeostasis that is not fully recapitulated by existing animal models. To reverse disease pathophysiology, we examined the effects of Nicotinamide Mononucleotide (NMN), a compound identified through a yeast-based drug repurposing screen for DHDDS-associated disease, both in patient-derived CBOs and clinically in six individuals enrolled in a 6-month, N-of-1 off-label treatment trial. Together, these findings provide mechanistic insight into the progressive neurological phenotype associated with heterozygous *DHDDS* variants and support the role of NMN as a potential therapeutic strategy for these severely affected patients.

## Results

### Patient Cohort

For this study, six patients harboring heterozygous *DHDDS* variants were included (**Table 1**).

**Table 1:**
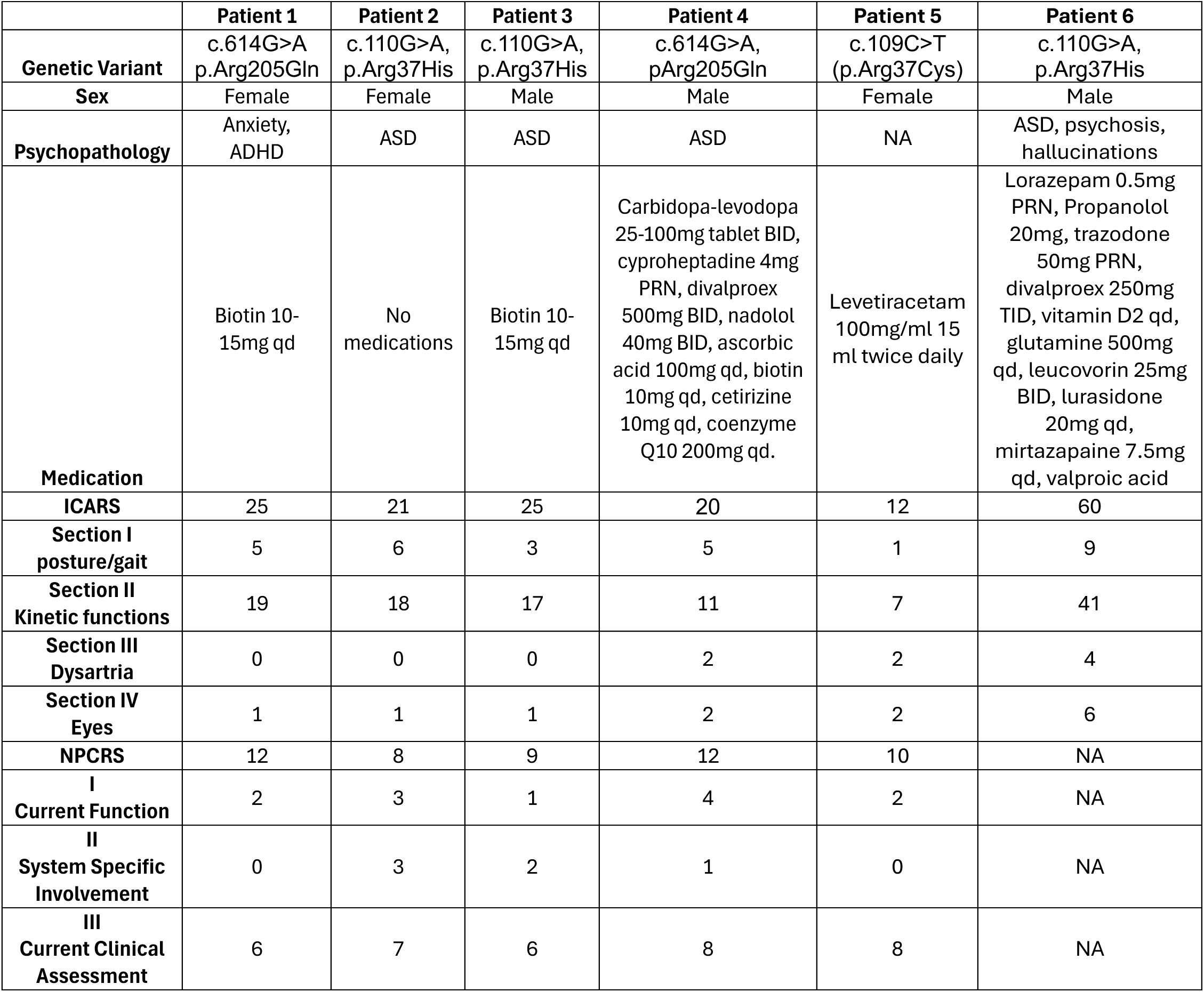
Overview of patients with heterozygous *DHDDS* variants.

Patient 1 presented with motor delay in infancy. She showed tremors and sleeping difficulties as a toddler and her first myoclonic jerks started to show as a child. Additionally, she has mild intellectual disability, anxiety and Attention-Deficit Hyperactivity Disorder (ADHD). Brain MRI did not show abnormalities. EEG showed generalized 4 Hz spike-wave activity. Patient 1 has a *de novo* heterozygous variant in *DHDDS:* c.614G>A, p.Arg205Gln. This variant was found to result in severely decreased DHDDS enzymatic activity in yeast.^9^

Patient 2 showed body tremors during infancy, together with hypotonia. Clumsiness and hypermobility were noted when she started walking. Myoclonus was first seen as a toddler. Her speech development seemed delayed as a toddler, although this concerned primarily her receptive language. She was diagnosed with an Autism Spectrum Disorder (ASD). Focusing, memory and information processing are challenging for her. Patient 2 has a heterozygous variant in *DHDDS*: c.110G>A, p.Arg37His. The p.Arg37His variant leads to severely reduced DHDDS activity both in yeast and mice.^9^

Patient 3 showed clumsiness and hypermobility when he started walking. Myoclonus and tremor were first seen as a child. He was diagnosed with ASD. His speech development is slightly delayed. Focusing, memory and information processing can be challenging. Patient 3 has a heterozygous variant in *DHDDS*: c.110G>A, p.Arg37His. The p.Arg37His variant leads to severely reduced DHDDS activity both in yeast and mice.^9^

Patient 4 has ASD, ataxia and mild intellectual disability. He was diagnosed with therapy resistant myoclonus epilepsy. EEG showed high amplitude (2-3 Hz), frontal predominant but relatively generalized, symmetrical irregular spike-and-wave burst activity. Brain MRI was normal. He has a c.614G>A, p.Arg205Gln variant in *DHDDS.* This variant was found to result in severely decreased DHDDS enzymatic activity in yeast.^9^

Patient 5 started to show seizures consisting of rhythmic twitching or fingers and arms as a child, together with dysarthria and tremor. She has mild intellectual disability. EEG showed mild diffuse slowing, generalized epileptiform discharges and paroxysmal focal slowing in the frontal regions. She has a c.109C>T, p.Arg37Cys variant in *DHDDS*. The p.Arg37Cys variant leads to severely reduced DHDDS activity both in yeast and in mice.^9^

Patient 6 shows seizures, sensorineural hearing loss, autism spectrum disorder, movement disorder including myoclonus, dystonia, ataxia. He has had psychosis and behavioral agitation. He has a heterozygous variant in *DHDDS*: c.110G>A, p.Arg37His. The p.Arg37His variant leads to severely reduced DHDDS activity both in yeast and mice.^9^

### Generation of patient-derived hiPSCs and transformation of hiPSCs to cortical brain organoids

Patient-derived fibroblasts were obtained from skin-punch biopsies from patient 1, 2 and 3 and were reprogrammed into human induces (hi) Primary Stem Cells (PSCs) using previously reported methods.^13^ To study neuronal development and function over time, we developed Cortical Brain Organoids (CBOs)^14,15^, containing multiple cell types, including glutaminergic– and GABAergic neurons, together with astrocytes.^15^ We found spheroid growth was comparable to healthy controls in the CBOs with heterozygous pathogenic *DHDDS* variants **(Figure 1A).** However, after 3-4 months, patient organoids started to shed dead cells and showed altered morphology compared to healthy controls, indicative of a degenerative phenotype **(Figure 1B).**

**Figure 1.**
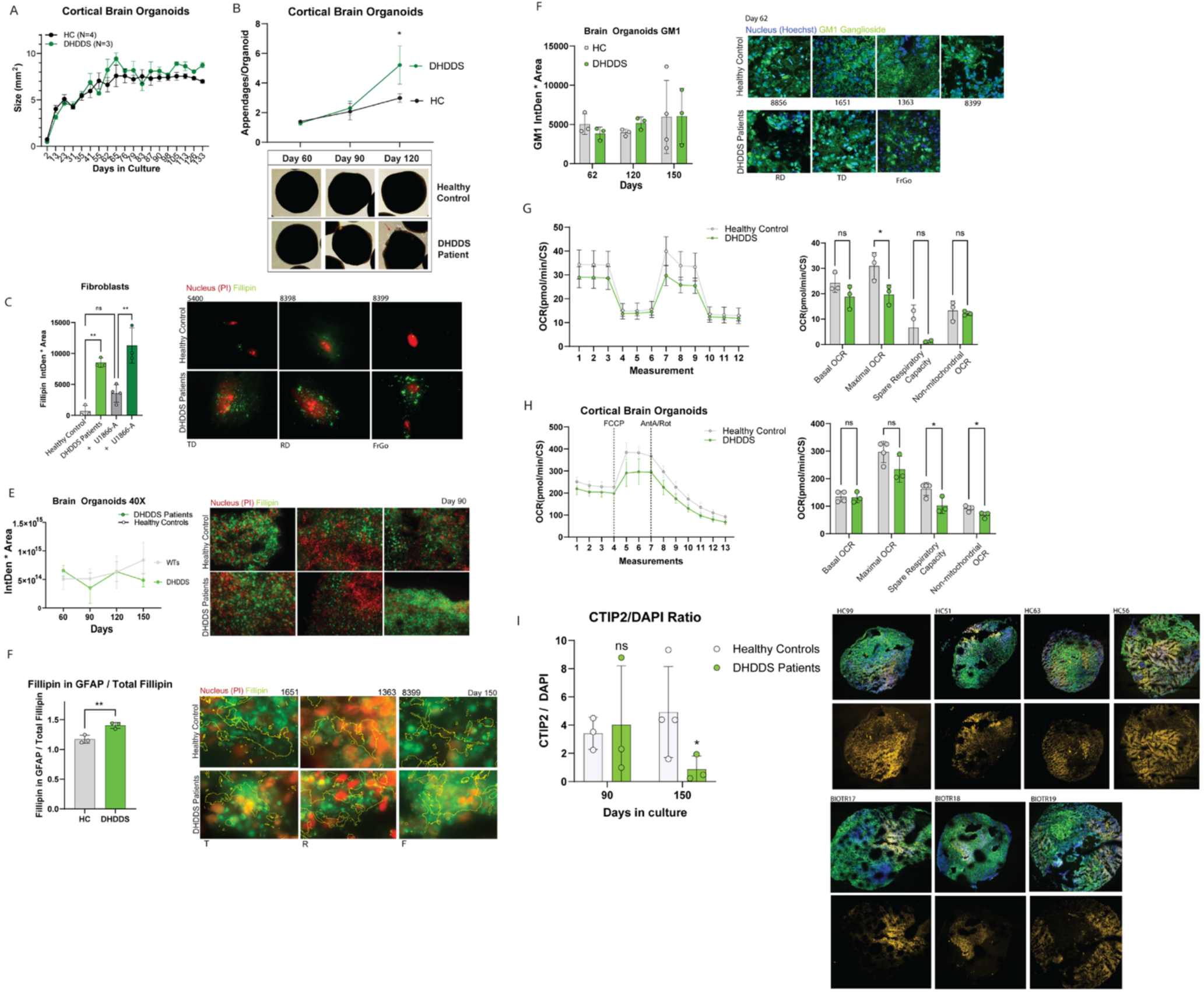
Cortical brain organoid characteristics and evaluation of GM1/ Cholesterol accumulation across cell types. A. Showing the total area (mm^2^) of cortical brain organoids over time. At least, three organoids per individual were quantified. Healthy controls (HC, N=4), patients with *DHDDS* variants (DHDDS, N=3). Line shows the average size of all individuals for the two groups ±SD. B. Showing the deterioration of organoids over time. Images of cortical brain organoids were assessed over time by quantifying how many appendages (>0.2 mm^2^) were present per organoid. Four images containing at least 2-3 organoids were quantified per individual. Healthy controls (N=4), patients with *DHDDS* variants (N=3). The lines show the mean number of appendages ±SEM. Two-way ANOVA followed by Šídák’s multiple comparison test. p<0.05*, p<0.01**, p<0.001***, p<0.0001****. C. Fillipin staining of fibroblasts of patients with *DHDDS* variants compared to healthy controls. Cholesterol abundance was quantified by calculating the integrated density (IntDen) values with ImageJ and multiplying this by the Fillipin total area. Healthy controls (N=4), patients with *DHDDS* variants (N=3). For each individual, at least eight different images were quantified. The bar graphs show the mean values of all individuals per group ±SD. Two-way ANOVA with Fisher’s LSD was used to calculate statistics. p<0.05*, p<0.01**, p<0.001***. D. Fillipin staining of cortical brain organoids of patients with *DHDDS* variants. Cholesterol abundance was quantified by calculating the integrated density (IntDen) values and multiplying those by the total area of the fillipin staining. Healthy controls (N=4), patients with *DHDDS* variants (N=3). For each individual, three 40X images of three different organoid slices were quantified (nine images in total). The lines show the mean of three individuals ±SD. Two-way ANOVA with Sídák as post hoc was used to calculate statistics. p>0.05 ns. E. Showing the ratio of mean intensity of the fillipin staining in astrocytes (GFAP+) compared to the total intensity of fillipin within the entire organoid. The bar graphs show the mean ratio of all individuals ±SD. Students’ t-test was used to calculate significance. p < 0.05*, p<0.001**. F. Showing the results of the Agilent XF96 mitochondrial stress test in fibroblasts of patients with *DHDDS* variants and healthy controls. The oxygen consumption rate was normalized to citrate synthase activity. Oligomycin (2.5 µM), FCCP (2 µM) and antimycin A/Rotenone (500 nM) were added during the assay. Four wells were analyzed for each of the individuals. Healthy controls (N=3) and patients with *DHDDS* variants (N=4). Both line graphs and bar graphs show the mean OCR for all individuals ±SEM. Two-way ANOVA followed by Šídák’s multiple comparison test was used to test for statistical significance. p<0.05*. G. Showing the results of the Agilent XF96 mitochondrial stress test in cortical brain organoids (at day 125) of patients with *DHDDS* variants and healthy controls. Oxygen consumption rate was normalized to citrate synthase activity. FCCP (2 µM) and antimycin A/Rotenone (500 nM) were added during the assay. Oligomycin was too large a molecule to penetrate the organoids and was removed from the assay. Four wells were analyzed for each of the individuals. Healthy controls (N=3) and patients with *DHDDS* variants (N=4). Both line graphs and bar graphs show the mean OCR for all individuals ±SEM. Two-way ANOVA followed by Dunnett’s multiple comparison test was used to test for statistical significance. p<0.05*. H. GM1 staining of cortical brain organoids at day 60,120 and 150. Healthy controls (N=4), patients with *DHDDS* variants (N=3). For each individual, three 60X images of three different organoid slices were quantified (nine images in total). The bar graphs show the Integrated density multiplied by the area of GM1. Statistics were calculated using a mixed effects model with Fisher’s LSD as post-hoc. p >0.05 ns. I. Showing the ratio of KI67+ total area over Doublecortin (DCX) area stained at day 60. For each individual, the average of at least 3 organoid slices/images was used. The bar graphs show the mean ratio of all individuals per group ±SD. Students T-test was used to calculate significance. p<0.05*. J. Showing the ratio of CTIP2+ cells compared to DAPI stained cells within cortical brain organoids at day 90 and 150. For each individual, the average of at least 3 organoid slices/images was used. The bar graphs show the mean ratio of all individuals per group ±SD. Mixed effect analysis was used to calculate statistics, with uncorrected Fisher’s LSD as post-hoc. p<0.05*.

### Cortical brain organoids of patients with pathogenic DHDDS variants show astrocyte-specific cholesterol accumulation

Previous work has shown substantial accumulation of unesterified cholesterol in patient-derived fibroblasts lines.^12^ We repeated the unesterified cholesterol staining in three patient-derived brain organoids with heterozygous *DHDDS* variants (patient 1, 2 and 3) and in three healthy controls **(Figure 1C)**. We found significant cholesterol accumulation in fibroblasts of patients with heterozygous *DHDDS* variants. As a positive control, we performed pre-treatment with U-18666A, a compound that impedes cholesterol trafficking. We found that this compound significantly induced cholesterol accumulation in healthy controls, indicating assay sensitivity **(Figure 1C).**

Despite cholesterol accumulation being observed in fibroblasts, recent work has found that cholesterol accumulation was absent from brain tissue of mice carrying the heterozygous *DHDDS* p.R37H variant.^11^ We assessed cholesterol accumulation in patient-derived CBOs, and found that cholesterol accumulation was not observed in whole organoid sections **(Figure 1D),** but localized to astrocytes specifically **(Figure 1E).** This concurs with the function of astrocytes, that are the main producents of cholesterol in the brain, while neurons can only take up the produced cholesterol derived from astrocytes.^16^ Of note, we did not observe unesterified cholesterol accumulation in patient-derived iPSCs **(Supplementary Figure 1A).**

Our findings indicate that cholesterol accumulation in individuals with pathogenic heterozygous *DHDDS* variants is restricted to astrocytes and absent in neurons, likely explaining why it was not detected in whole-brain mouse or human organoid sections. We found no evidence of GM1 accumulation in either neurons or astrocytes in cortical brain organoids.

### Fibroblasts and cortical brain organoids of patients with heterozygous DHDDS variants show mitochondrial dysfunction

Next, we assessed mitochondrial function using Seahorse respirometry. In patient-derived fibroblasts, we found a significant reduction of maximum respiratory capacity and a reduction of spare respiratory capacity **(Figure 1F)**. Similarly, in patient-derived CBOs, we observed decreased maximum– and significantly decreased spare respiratory capacity **(Figure 1G).** We verified whether mitochondrial morphological changes were at associated with mitochondrial dysfunction, but mitochondrial morphology was similar between healthy controls and patient-derived fibroblasts of individuals with heterozygous *DHDDS* variants **(Supplementary Figure 1C).**

### Cortical brain organoids of patients with pathogenic DHDDS variants do not show GM1 accumulation

Recently, increased GM1 accumulation has been observed in fibroblasts of patients with *DHDDS* variants when compared to one healthy control.^12^ However, we did not observe ganglioside (GM1) accumulation in whole-organoid section of DHDDS patient-derived organoids **(Figure 1H)**. We also assessed GM1 accumulation in astrocytes specifically, where GM1 accumulation was similarly not observed **(Supplementary Figure 1B)**.

### Cortical brain organoids of patients with DHDDS variants show decreased levels of KI67+ neurons early in development, and loss of deep-layer neurons during aging

To study the development of CBOs in patients with *DHDDS* variants, we assessed several developmental markers at different timepoints. We found that the early developmental marker KI67 was significantly decreased by day 60 in patients with *DHDDS* variants, indicating either a decreased level of developing neurons or maturation that is finished earlier within development **(Figure 1I).** The level of doublecortin was comparable between patients and healthy controls on day 60 **(Figure 1I).**

In patient-derived CBOs with primary mitochondrial dysfunction, there is significant apoptosis of deep-layer neurons (CTIP2+), as these neurons have high metabolic demands making them vulnerable to apoptosis and oxidative stress.^17^ Since patients with *DHDDS* variants similarly show mitochondrial dysfunction, we verified if their CBOs showed decreased numbers of deep layer neurons. While the overall level of neurons (MAP2+) was similar between healthy controls and patients (data not shown), the number of deep layer neurons (CTIP2+) was similar at day 90, but significantly decreased at day 150 **(Figure 1J)**. These data suggest that deep-layer neurons develop normally in early development, but degenerate during aging, similar to patients with primary mitochondrial dysfunction.

### Temporal proteomic profiling reveals altered neurodevelopment, axon regeneration, and lipid metabolism in DHDDS cortical organoids

To assess which pathways were significantly affected in cortical organoids harboring *DHDDS* variants, we next performed proteomic analysis on cultures maintained for either 60 days, 90 days, and 120 days **(Supplementary Table 1)**. The rationale for these time points was to include developmental stages both around and after the gliogenic switch (∼day 100). The three timepoints showed distinct clustering on Uniform Manifold Approximation (UMAP) (**Figure 2A).** Several proteins involved in lipid metabolism were significantly altered across different timepoints in patients with heterozygous *DHDDS* variants, including a downregulation of DDHD2 and PIP4K2A, and an upregulation of PAFAH1B2 **(Figure 2B).** UBR2, a ubiquitin protein ligase E3 that is involved in protein degradation and dampening ER stress^18^, was upregulated at day 60 and day 90 **(Figure 2B).**

**Figure 2:**
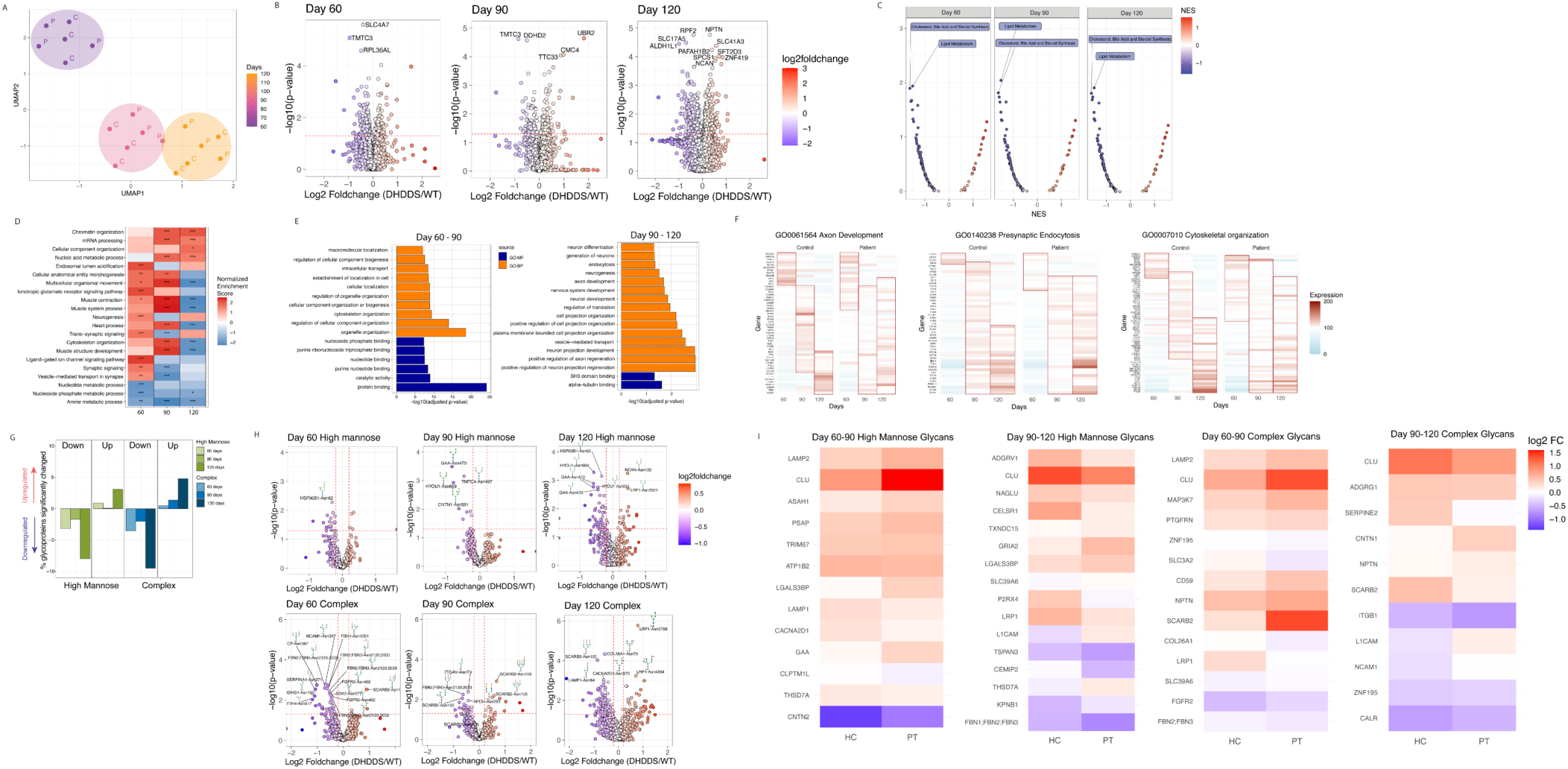
Proteomic results of cortical brain organoids of patients with *DHDDS* variants across developmental stages. A. Uniform manifold approximation (UMAP) showing distinct clusters for each of the developmental stages tested in cortical brain organoids: day 60, day 90 and day 120. N-neighbors = 9 and minimal distance = 0.01. For proteomics, 4 healthy control and 3 patient samples were included for each of the three timepoints. B. Volcano plots highlighting the differentially expressed proteins (p-value <0.01 and log2foldchange value >0.2, mixed model analysis with Turkey’s HSD test) for the three developmental stages (day 60, day 90 and day 120). The X-axis shows the log2 foldchange of the protein expression of individuals with *DHDDS* variants over healthy controls, while the Y-axis shows the –log10(p-value). C. Showing the Gene Set Enrichment results for the MitoCarta gene sets specifically. The x-axis shows the Normalized Enrichment Score (NES), the y-axis shows the –log10(nominal p-value). D. Gene set enrichment analysis (GSEA) for the three different timepoints assessed (day 60, day 90 and day 120). Proteins were sorted based on log2foldchange and pre-ranked GSEA was performed. The top-20 pathways with the lowest adjusted p-values (FDR adjusted) were selected for each of the three timepoints. For better visualization, ontologies belonging to the same hierarchy were merged using the EMBL-EBI database. The mean p-values and NES are shown. p<0.05*, p<0.01**, p<0.001***, p<0.0001****. E. Pathway analysis of the differential protein expression over time. First, the difference in expression between timepoint 90-60 days and 90-120 days was calculated (delta) for both healthy controls and patients. Next, we tested which differences (deltas) were significantly different between healthy controls and patients with *DHDDS* variants (p-value <0.05). These proteins were subsequently assessed using overrepresentation analysis with g:profiler (version e113_eg59_p19_f6a03c19).^38^ F. Showing the expression of individual proteins of the significant GO ontologies from Figure 2E at different time points. As can be seen, the expression of proteins is significantly altered in patients with *DHDDS* variants at the different time points for proteins involved in axon development, presynaptic endocytosis and cytoskeletal organization. G. Showing the percentage of glycan species that were significantly up– or downregulated at three distinct time points (day 60, day 90 and day 120). Cutoff values for significance were p-value 0.05 (Linear Mixed Model analysis with Turkey’s HSD) and log2 foldchange of < –0.2 or > 0.2. H. Volcano plots highlighting the differentially expressed glycopeptides (p-value <0.01 and log2foldchange value >0.2, mixed model analysis with Turkey’s HSD test) for the three developmental stages (day 60, day 90 and day 120). The X-axis shows the log2 foldchange of the glycopeptide expression of individuals with *DHDDS* variants over healthy controls, while the Y-axis shows the –log10(p-value). The upper three volcano plots show high-mannose glycan species, while below, complex/hybrid glycan species are shown. I. Showing all proteins whose glycosylation changes significantly over time (p<0.01 and log2foldchange < –0.5 or >0.5). The average of all glycosylation sites and species in healthy controls was calculated and divided by the average of patients, resulting in log2foldchange values. The majority of proteins kept similar glycosylation levels over time, both in healthy controls and in patients.

Considering the mitochondrial dysfunction observed in CBOs of patients, we also assessed the expression of mitochondrial gene sets specifically. Intriguingly, we observed that out of all MitoCarta Gene Sets, “Lipid Metabolism” and “Cholesterol and Steroid synthesis” were significantly downregulated across all developmental timepoints **(Figure 2C)**. This concurs with observations in a mouse model with a heterozygous p.R37H *DHDDS* variant, showing altered abundance of triglycerides and phospholipids in whole brain lysates.^11^ In addition, pathway analysis showed significant upregulation of mRNA processing and nucleic acid metabolism across all timepoints in patients with *DHDDS* variants **(Figure 2D).** Intriguingly, neurogenesis, synaptic signaling and vesicle-mediated transport in synapses were upregulated at early timepoints (day 60) but downregulated later in development (day 90-120) **(Figure 2D).** Protein-, nucleotide– and nucleoside metabolism were downregulated across all timepoints in patients with *DHDDS* variants **(Figure 2D).**

Next, we studied how protein expression varied throughout development. To this end, we studied the difference in protein expression between day 60 – day 90, and day 90 – day 120. We compared these differences between healthy controls and patients with *DHDDS* variants and found several pathways showing altered protein expression in early development (day 60 – 90), including cytoskeletal organization and intracellular transport **(Figure 2E).** Interestingly, proteins whose expression differed significantly between day 90 and 120 were enriched in pathways related to neuronal development and axon regeneration **(Figure 2E,F)**. Vesicle-mediated transport was altered in early development (day 60-90) and around the gliogenic switch (between day 90 and 120) **(Figure 2E,F).**

Thus, our proteomics results suggest that *DHDDS* variants disrupt several pathways, including lipid and mRNA/nucleic acid metabolism. In addition, the temporal dynamics of key neurodevelopmental, cytoskeletal and vesicle-trafficking pathways in CBOs of patients with *DHDDS* variants were altered.

### Glycoproteomics results-show glycosylation abnormalities that worsen over time in patients with DHDDS variants

In addition to proteomics, we also performed glycoproteomics across three developmental time points in the CBOs of patients with *DHDDS* variants **(Supplementary Table 2)**. **Figure 2G** shows the significantly changing glycopeptides in cortical brain organoids of patients with *DHDDS* variants across timepoints. The downregulation of both high-mannose and complex/hybrid glycans in patients was strongest later in development, around 120 days of age. Several proteins showed aberrant glycosylation across multiple timepoints, including HSP90B1, GAA, HYOU1, SCARB2 and FBN2/FBN3 **(Figure 2H).** We also assessed whether specific glycopeptides showed differences in up– or downregulation across time between healthy controls and patients. To this aim, we studied significantly changing glycopeptides between day 60 – day 90 and day 90 – day 120. Most glycopeptides became more– or less glycosylated in the same manner across healthy controls and patients with time **(Figure 2I)**. Only SLC39A6, L1CAM, THSD7A and SERPINE2 were differentially glycosylated in patients between timepoint 90 and 120.

Interestingly, there were several proteins aberrantly glycosylated that are involved in lipid metabolism or lysosomal function. One example is APOH that was differentially glycosylated at day 90, which is known to bind to triglycerides and phospholipids^19^. In concurrence, the abundance of phospholipids and triglycerides was significantly altered in cortical brain organoids heterozygous *DHDDS* variant at day 90 **(Figure 3D)**. N-glycosylation helps to maintain the closed circular conformation of APOH and aberrant glycosylation promotes binding to negatively charged phospholipids.^20^ We also observed that SCARB2/LIMP2 was aberrantly glycosylated across developmental timepoints. LIMP2 enables the transport of lipids like phospholipids, sphingosine, and cholesterol from the lysosomal lumen to the membrane.^21^ Thus, altered glycosylation of LIMP2 might affect trafficking of these molecules.

**Figure 3:**
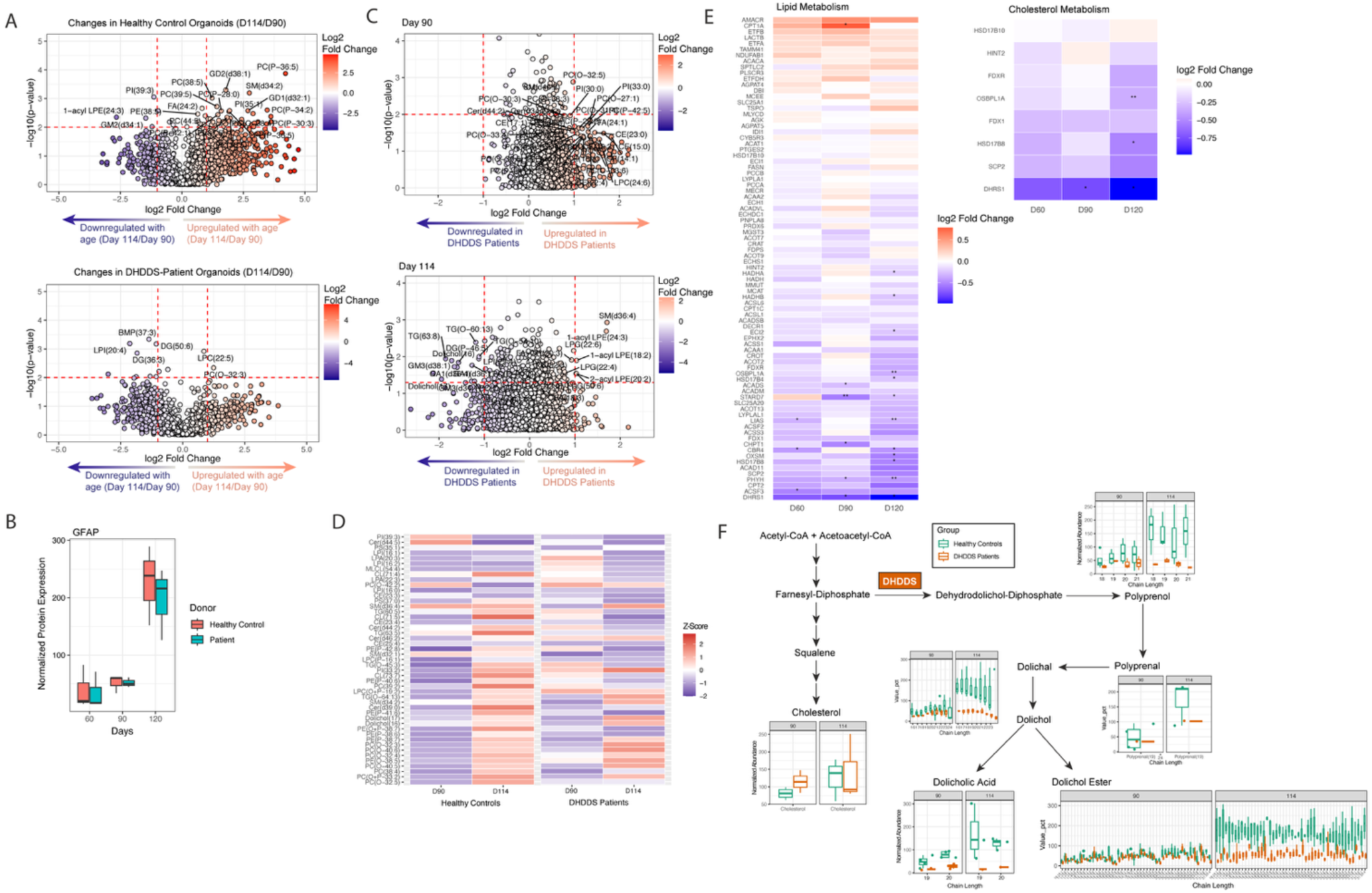
Lipidomic changes in patients with *DHDDS* variants reveals progressive dolichol depletion with advancing organoid age. A. Volcano plot showing the changes in lipid species in healthy control (N=3) and patient-derived cortical brain organoids (N=3) with advancing organoid age (day 90 versus day 114). Paired t-tests were used to calculate significance. B. Showing normalized GFAP expression from proteomics data at three timepoints (day 60, day 90 and day 120) in healthy controls and DHDDS patients. The boxplots show the median expression value for the individuals ±25^th^-75^th^ percentile and extend to 1.5*IQR. C. Volcano plot showing the log2foldchange of lipid species of patients with *DHDDS* variants (N=3) over healthy controls (N=3) at day 90 and day 114. Linear mixed models with Turkey’s HSD test were used to calculate significance. D. The Z-scores of the lipid species that significantly increased or decreased (Welsch t-test p<0.01) on day 114 compared to day 90 are shown. Z-scores were calculated per individual. E. Showing protein expression over time (log2 foldchange of DHDDS patients over healthy controls) of proteins involved in lipid metabolism and cholesterol metabolism. The stars reflect the p-value that was calculated with mixed model analysis and Turkey’s HSD. p<0.05*, p<0.01**, p<0.001***. F. Showing the abundance of different lipid species involved in dolichol metabolism in patients with *DHDDS* variants and healthy controls at day 90 and day 114. All values in patients were normalized to healthy control values. Polyprenol, polyprenal, dolichoic acid and dolichol esters were annotated based on clustering logic (distinct m/z differences and retention time correlations characteristic to a homologous series). MS/MS fragmentation was not performed to confirm these structures.

In conclusion, heterozygous *DHDDS* variants cause unique glycosylation defects in LIMP2 and APOH, which may contribute to altered lysosomal lipid trafficking and phospholipid binding.

### Lipidomics reveals progressive dolichol depletion and altered phospholipid metabolism in patients with DHDDS variants

Next, we assessed lipid metabolism alterations in CBOs of patients with *DHDDS* variants before and after the gliogenic switch (day 90 and 114) **(Supplementary Table 3).** Intriguingly, there was a massive upregulation of phosphatidylcholine ether lipids (PC-O and PC-P) species after the emergence of astrocytes in healthy control organoids **(Figure 3A)**. This might be due to the higher fatty oxidation levels in astrocytes, that in turn leads to increased production of phosphatidylcholine ether lipids.^22,23^ However, this massive upregulation of phosphatidylcholine ether species between day 90 and day 114 was not observed in patient-derived cortical brain organoids. This was not due to earlier or later maturation of astrocytes, since they developed at similar rates **(Figure 3B).** Intriguingly, the majority of lipids were upregulated at day 90 in DHDDS patients compared to healthy controls **(Figure 3C)**. At day 114, lipid expression was comparable between DHDDS patients and healthy controls, however, the downregulation of dolichol species was the most differentiating feature observed **(Figure 3C)**. There were numerous lipid species that showed a different developmental trajectory in DHDDS patients over time, including ceramides, cardiolipins, cholesterylesters, phosphatidylethanolamines, phosphatidylcholines and phosphatidylcholine ethers **(Figure 3D).** This might correspond with altered expression of several enzymes involved in lipid– and cholesterol metabolism that was observed with proteomics **(Figure 3E)**. Interestingly, dolichol levels and dolichol-related molecules were deceased in patients with *DHDDS* variants **(Figure 3F)**. This depletion appeared to be progressive, as dolichol, polyprenol, polyprenal, dolichoic acid and dolichol ester levels increased in healthy controls with age, but remained low in patients over time **(Figure 3F).**

Thus, lipidomic analysis of CBOs revealed a pronounced astrocyte-associated upregulation of phosphatidylcholine species after the gliogenic switch in healthy controls that was absent in DHDDS patient-derived organoids, which instead showed early lipid alterations and a progressive, age-dependent depletion of dolichol species.

*NAD+ related molecules improve growth and survival of a yeast DHDDS model* Because *cis*-PTase–dependent dolichol biosynthesis is conserved from yeast to humans, we performed a small-molecule drug screen in a previously described humanized yeast model expressing the pathogenic p.Arg205Gln (R205Q) *DHDDS* variant to identify compounds capable of rescuing disease-associated growth defects.^9^ In this model, the endogenous cis-PTase subunits (rer2, srt1, and nus1) are deleted and initially complemented by a URA3-marked plasmid expressing *Giardia lamblia* cis-PT. Following counterselection on 5-fluoroorotic acid (FOA), growth becomes dependent on co-expression of human DHDDS and NgBR, rendering yeast viability contingent on functional human cis-PTase activity.^9^

Consistent with prior reports^9^, p.R205Q-expressing yeast impaired growth relative to the wild-type control (**Supplementary Figure 2).** Screening of an ∼8,400-bioactive small-molecule library identified several compounds that partially rescued this growth defect **(Figure 4A).** Intriguingly, NADP and NADPH emerged among the top rescuers, and dose-response experiments confirmed that NADPH enhanced mutant DHDDS-deficient yeast growth at higher concentrations **(Figure 4B).**

**Figure 4:**
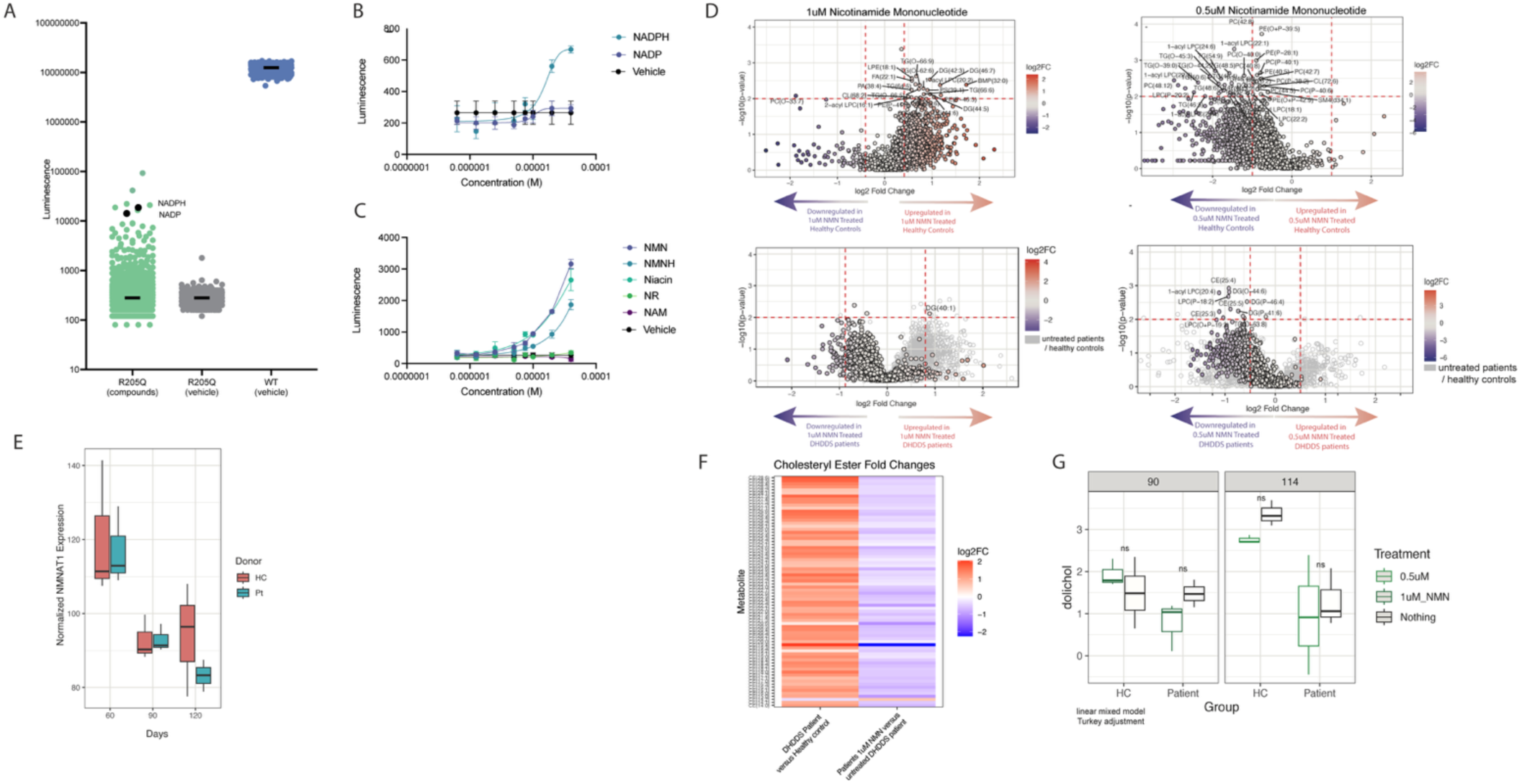
The identification of Nicotinamide Mononucleotide (NMN) as a potential treatment target and its effects on lipid profiles in cortical brain organoids. A. Results of a high-throughput screen of 8,387 bioactive compounds in a humanized yeast model expressing the DHDDS p.R205Q variant. R205Q mutant yeast treated with DMSO vehicle are shown in grey. Yeast expressing wild-type human DHDDS and NgBR (WT) are shown in blue. R205Q mutant yeast treated with compounds from the screening library are shown in green. Each point represents ATP-based luminescence measured following treatment with individual compounds at 20 µM. NADPH and NADP are highlighted as black points. Horizontal black bars indicate the median luminescence for each condition. B. Dose–response effects of NADP and NADPH on the growth of DHDDS p.R205Q-expressing yeast. Mutant yeast were treated with increasing concentrations of NADP or NADPH or with DMSO vehicle, and growth was assessed by ATP-based luminescence. Data represent mean ± SD of technical replicates (n = 3 wells per concentration; n = 16 wells vehicle). C. Dose–response effects of NAD⁺ precursors in DHDDS p.R205Q-expressing yeast. Mutant yeast were treated with increasing concentrations of niacin, nicotinamide mononucleotide (NMN), dihydronicotinamide mononucleotide (NMNH), nicotinamide riboside (NR), or nicotinamide (NAM), or with DMSO vehicle, and growth was assessed by ATP-based luminescence. Data represent mean ± SD of technical replicates (n=3 wells per concentration; n=16 wells vehicle) Showing the effects of 2-week 0.5µM and 1µM NMN treatment on lipid species in cortical brain organoids before the gliogenic switch (day 90, 1µM NMN left) and after (day 114, 0.5µM NMN, right). Statistics were calculated using Welsch t-test. D. Showing the protein expression of NMNAT1 over time in cortical brain organoids, the protein that converts NMN to NAD+ in the nucleus. The boxplot shows the median expression value and extends to the 25^th^-75^th^ percentile. The lines extend to 1.5*IQR. E. Showing the effect of NMN treatment on cholesteryl esters on day 90. On the left, the log2foldchange of patients over healthy controls is shown of cholesteryl esters, while on the right the log2foldchange (Log2FC) of treated patients is shown over untreated patients. F. Showing the median dolichol expression of all chain lengths at the different time points assessed and the effects of NMN. The boxplot shows the median expression value and extends to the 25^th^-75^th^ percentile. The lines extend to 1.5*IQR. Statistics were calculated using mixed model analysis with Turkeys HSD as post-hoc.

Given the central role of NAD(P)(H) in cellular metabolism, we next evaluated whether other NAD⁺-related metabolites could similarly improve yeast growth. We found that niacin, nicotinamide mononucleotide (NMN), and dihydronicotinamide mononucleotide (NMNH) significantly rescued mutant yeast growth, whereas nicotinamide riboside (NR) and nicotinamide (NAM) showed no effect **(Figure 4C).** Subsequently, we tested whether these NAD+ precursors were able to rescue the disease phenotypes in CBOs of individuals with heterozygous *DHDDS* variants.

### Metabolic changes in cortical brain organoids after NMN treatment

We assessed the lipidomic changes in CBOs before and after NMN treatment for 2 weeks at different concentrations (0.5 and 1 µM). We found that 1µM NMN treatment led to an upregulation of lipid species at day 90 in healthy controls, while it had the opposite effect on lipid species in patient with *DHDDS* variants **(Figure 4D).** At day 114, 0.5 µM NMN decreased most lipid abundances both in healthy controls and in patients with *DHDDS* variants **(Figure 4D)**. The emergence of astrocytes might explain how NMN first downregulates lipids in patients and upregulates lipids in healthy controls, while at day 114 downregulates lipids in both healthy controls and patients. A potential explanation for the difference in NMN effects at day 90 and day 114 in healthy controls could be the decline in NMNAT1 levels with advancing age **(Figure 4E)**. NMN significantly reduced cholesteryl esters in patient-derived cortical organoids **(Figure 4F)**. NMN did not significantly affect dolichol levels in either healthy controls or patients **(Figure 4G).**

At day 90, cholesteryl esters were significantly increased in patients with *DHDDS* variants, and NMN was able to significantly reduce these cholesteryl esters **(Figure 4F)**. Cholesteryl esters accumulate in brain cells of patients with Alzheimer’s disease.^24^ NMN did not affect dolichol levels significantly in either healthy controls or patients **(Figure 4G).**

### Nicotinamide mononucleotide rescues the mitochondrial phenotype in patient-derived organoids of patients with heterozygous DHDDS variants

Next, we verified the effects of NMN using Seahorse respirometry. We found that NMN led to significantly enhanced maximal and spare respiratory capacity rates in DHDDS organoids, indicating improved mitochondrial function **(Figure 5A/B)**. This improvement was observed for both the 0.5μM and 1μM concentration.

**Figure 5:**
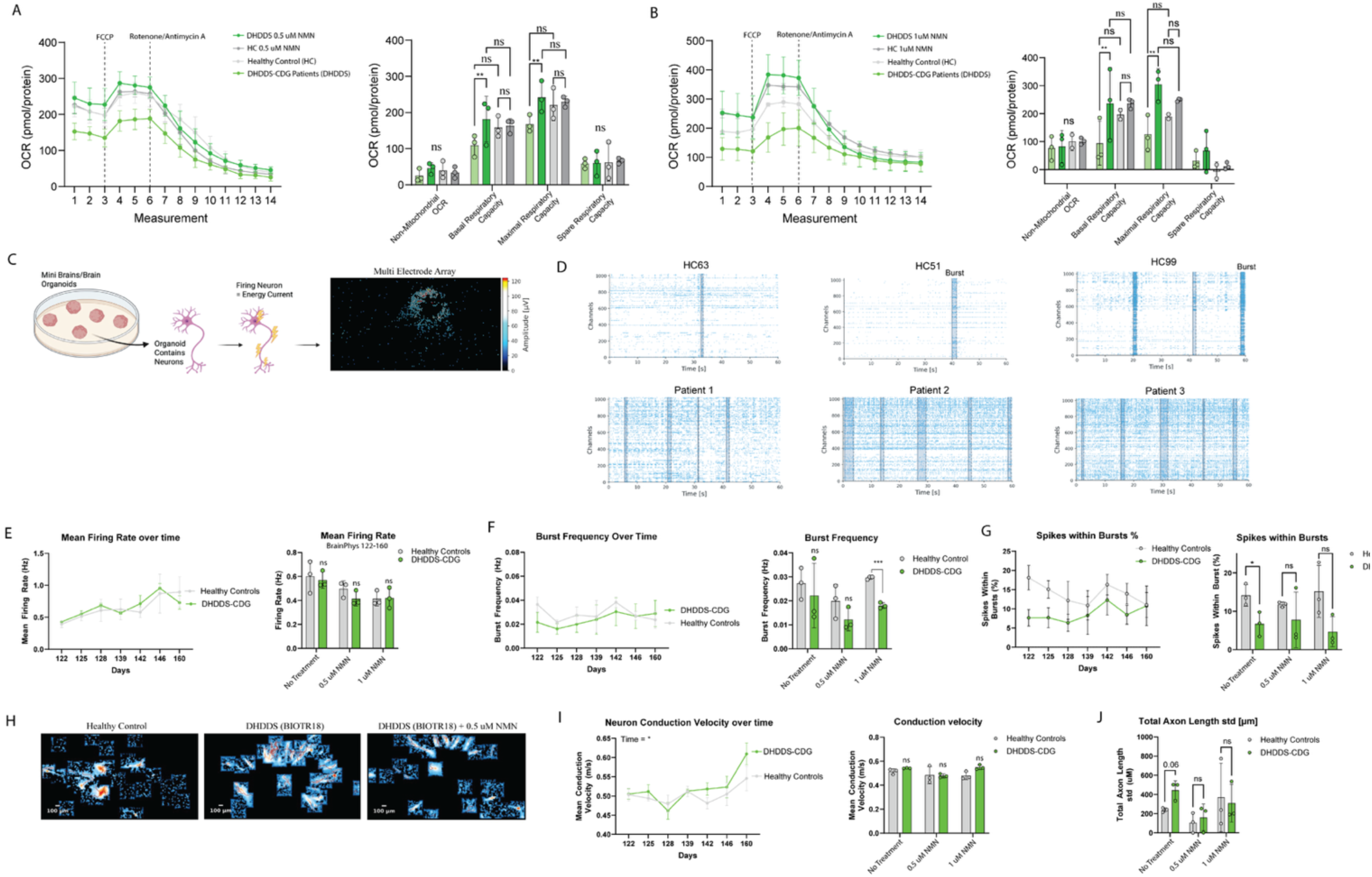
Treatment effects of Nicotinamide Mononucleotide (NMN) on mitochondrial function and electrophysiological activity in cortical brain organoids. A. Seahorse respirometry results before and after treatment with 0.5µM NMN in cortical brain organoids. Since oligomycin does not penetrate the organoids, this treatment was left out. On the left, the mean OCR ±SEM is shown before and after the injection with different compounds (FCCP, Rotenone/Antimycin A, same protocol as Figure 1H). On the right, bar graphs reflecting the mean ±SD of the different measures (basal-, maximal-, spare respiratory capacity and non-mitochondrial respiration) are shown. Mixed effect models with Fisher LSD analysis were used to calculate statistics. p <0.05*, p<0.01**. B. Seahorse respirometry results before and after treatment with 1µM NMN in cortical brain organoids. Since oligomycin does not penetrate the organoids, this treatment was left out. On the left, the mean OCR ±SEM is shown before and after the injection with different compounds (FCCP, Rotenone/Antimycin A, same protocol as Figure 1H). On the right, bar graphs reflecting the mean ±SD of the different measures (basal-, maximal-, spare respiratory capacity and non-mitochondrial respiration) are shown. Mixed-effect models with Fisher LSD were used to calculate statistics. p <0.05*, p<0.01**. C. Graphical representation of Multi Electrode Array (MEA) workflow. D. Representative images of MEA results. The black dashed squares reflect network bursts. E. Showing the mean firing rate (averaged across all three individuals) ±SEM over time in aging organoids. There were no significant differences between healthy controls and patients with *DHDDS* variants (two-way ANOVA). The bar graph shows the results of NMN treatment on Mean Firing Rate. To create the bar graphs, different time points (days) were averaged for each of the individuals. The bar graphs show mean ±SD for the three different individuals. There were no significant differences between untreated and treated organoids for healthy controls or patients with *DHDDS* variants (two-way ANOVA). F. Showing the burst frequency over time in aging organoids. There were no significant differences between healthy controls and patients with *DHDDS* variants (two-way ANOVA). The bar graph shows the results of NMN treatment on Burst Frequency. While there was a trend towards decreased burst frequency in the patients, only the 1µM condition untreated and treated organoids for healthy controls or patients with *DHDDS* variants was significantly different (two-way ANOVA). To create the bar graphs, different time points (days) were averaged for each of the individuals. The bar graphs show mean ±SD for the three different individuals. p <0.05*, p<0.01**, p<0.001***. G. Showing the number of spikes (action potentials) that occur within or outside network bursts over time in aging organoids. In mature organoids, the majority of spikes are organized within network bursts. The number of spikes within bursts was significantly lower in patients with *DHDDS* variants (two-way ANOVA). The bar graph shows the results of NMN treatment on the mean number of spikes that occurred within bursts. There was a significant difference between healthy controls and patients with *DHDDS* variants in the untreated condition (two-way ANOVA). This became unsignificant after treatment. To create the bar graphs, different time points (days) were averaged for each of the individuals. The bar graphs show mean ±SD for the three different individuals. p <0.05*. H. Representative images of the axon tracking assay assessed using MEA. The blue colors refer to electric activity being registered. The red lines indicate propagation within individual neurons. I. Showing the speed of neuron conduction velocity (meter/second) in aging organoids. The neuron conduction velocity increased significantly over time. The bar graph shows the results of NMN treatment on the neuron conduction velocity. There were no significant differences between healthy controls and individuals with *DHDDS* variants with and without NMN treatment. To create the bar graphs, different time points (days) were averaged for each of the individuals. The bar graphs show mean ±SD for the three different individuals. p <0.05*. J. Showing the total axon length in aging organoids. The bar graph shows the results of NMN treatment on the neuron conduction velocity. There were no significant differences between healthy controls and individuals with *DHDDS* variants with and without NMN treatment. To create the bar graphs, different time points (days) were averaged for each of the individuals. The bar graphs show mean ±SD for the three different individuals. p <0.05*.

Neurons produce individual action potentials, so-called “spikes”. Upon maturing, organoids start to produce synchronous, spontaneous electrical activity rather than individual spikes, called “Network Bursts” **(Figure 5C,D).** These signatures can be captured using Multi Electrode Array (MEA). As such, we compared untreated and treated organoids for two weeks with 0.5μM NMN. Later in development, we increased the dosage from 0.5μM to 1μM of NMN, to see if phenotypes would improve even further.

We found the average spike rate and burst frequency were not significantly different in patient-derived organoids and healthy controls, which did not change with NMN treatment **(Figure 5E, F)**. In mature organoids, spikes are usually produced within a network burst and activity should be limited outside of these bursts. However, we found the number of spikes outside of network bursts seemed increased in organoids of patients with *DHDDS* variants, suggesting a degree of asynchronicity, especially at early timepoints **(Figure 5G)**. Two weeks of 0.5μM NMN treatment normalized the percentage of spikes within bursts **(Figure 5G).** 1μM of NMN treatment do not appear to normalize the number of spikes within bursts, however, this treatment was applied later in development (day 144-160), when the synchronicity in patients was already more matured.

The MEA system also tracks the propagation of action potentials through individual neurons within the organoid **(Figure 5H).** We found the average conduction velocity of action potentials was not significantly different in patients with *DHDDS* variants and healthy controls, with and without NMN treatment **(Figure 5I).** However, the variation in axon length seemed higher in patients with *DHDDS* variants (p=0.06). This increased variation normalized upon NMN treatment **(Figure 5J).**

Together, these results indicate that NMN partially rescues mitochondrial dysfunction in patients with *DHDDS* variants, highlighting its potential as a therapeutic candidate. We observed some mild electrophysiological phenotypes, which could be improved under NMN.

### Clinical improvements during oral NMN treatment in 6 patients in a series of N-of-1 off-label trial

The yeast model drug screen results prompted the patients to initiate 250 mg/day oral NMN supplementation individually **(Table 2)**. Since NMN is a nutritional supplement, it can be bought without prescription. Six patients without severe intellectual disability who were able to complete ICARS assessments were followed over a period of 6-months after initiation of 250 mg NMN orally. Their clinical details are listed in **Table 1**. Their ages ranged from 10 to 19 years old. We enrolled two males and four females, with ICARS scores at baseline ranging from 12 to 60 (out of 100).

**Table 2:** ICARS scores after 6 months of oral Nicotinamide Mononucleotide treatment.

|  | Patient 1 | Patient 2 | Patient 3 | Patient 4 | Patient 5 | Patient 6 |
| --- | --- | --- | --- | --- | --- | --- |
| **Genetic Variant** | c.614G>A, p.Arg205Gln | c.110G>A, p.Arg37His | c.110G>A, p.Arg37His | c.614G>A, p.Arg205Gln | c.109C>T (p.Arg37Cys) | c.110G>A, p.Arg37His |
| **Sex** | Female | Female | Male | Male | Female | Male |
| **NMN Treatment Duration (months)** | 14 | 10 | 10 | 9 | 6 | 6 |
| **ICARS at Baseline** | 25 | 21 | 25 | 20 | 12 | 60 |
| ICARS after 6 months of 250mg NMN | 19 | 18 | 18 | 12 | 7 | 38 |
| ICARS Section I: Posture Gait | 2 | 4 | 3 | 3 | 1 | 4 |
| ICARS Section II: Kinetic Function | 14 | 14 | 15 | 7 | 4 | 31 |
| ICARS Section III: Dysarthria | 0 | 0 | 0 | 2 | 1 | 2 |
| ICARS Section IV: Eyes | 3 | 0 | 0 | 1 | 1 | 1 |

During the trial period, ICARS was assessed at baseline and after 6 months of oral NMN supplementation. Additionally, patients recorded videos at home of selected ICARS tests, including tandem walking and spiral drawing.

To quantify ataxia, we assessed videos showing the deviation from the midline of tandem walking based on arm– or leg movement **(Figure 6A/B).** For 5 out of 6 patients with heterozygous *DHDDS* variants, tandem walking showed significant deviation from the midline compared to healthy, age-matched children. One patient showed tandem walking with arm– and leg deviation within the normal range, however, his movements were very rigid and slowed **(Figure 6B).** Deviation from the midline was quantified before and after 6 months of NMN supplementation **(Figure 6C).** All patients improved their tandem walk significantly **(Figure 6D).** One patient (patient 3) remained stable during the 6 months of treatment, however, his deviation from the midline was already similar at baseline to aged-matched healthy controls.

**Figure 6:**
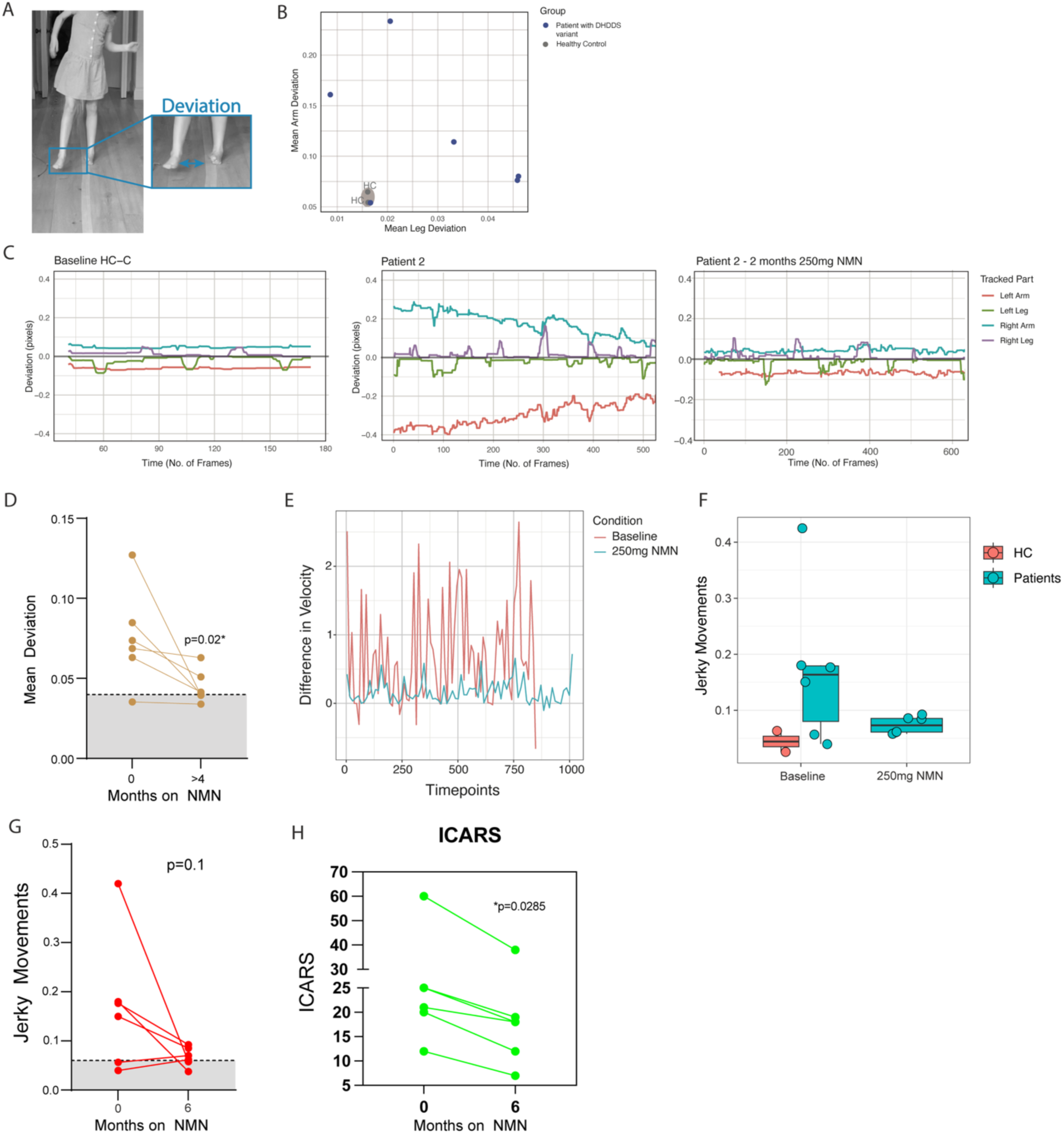
Treatment effects of Nicotinamide Mononucleotide (NMN) in 6 patients on an off-label N-of-1 trial. A. Graphical representation of how deviation of patients’ hand and feet from the midline was quantified. B. Showing the mean deviation of the arms (y-axis) and legs (x-axis) from a midline drawn on the ground during video assessments of two healthy pediatric controls (in grey) and patients (blue). C. Showing the mean deviation of the arms and legs from a midline drawn on the ground during video assessments of two healthy pediatric controls (in grey) and patient 2 before NMN treatment and after NMN treatment (right two graphs, respectively) performing a tandem walk. D. Showing the mean deviation of arms and legs at baseline (timepoint 0) and after 4 or 6 months of treatment (timepoint >4 months). Paired t-test was used to calculate significance. p>0.05 ns, p<0.05 *. E. Showing the smoothness of drawing the Archimedes spiral in patients with *DHDDS* variants and healthy controls. To quantify smoothness, the difference in velocity during drawing was quantified. The standard deviation of the velocity differences is shown on the Y-axis (“Jerky Movements”). The difference between the groups was non-significant (Welsch t-test). p>0.05 ns. F. Showing the difference in velocity (Y-axis) over time (X-axis) during the drawing of the Archimedes spiral of patient 3 at baseline and after 6 months of NMN. G. Showing the smoothness of drawing the Archimedes spiral in patients with *DHDDS* variants and healthy controls at baseline and after 6 months of NMN treatment. To quantify smoothness, the difference in velocity during drawing was quantified. The standard deviation of the velocity differences is shown on the Y-axis (“Jerky Movements”). Paired t-test was used to calculate significance. p>0.05 ns. H. Showing the ICARS scores before and after NMN treatment with 250mg. Paired t-tests were used to calculate significance. p>0.05 ns, p<0.05 *, p<0.001**.

We also quantified the drawing of the Archimedes spiral before and after NMN treatment. Interestingly, the deviation of the drawn spiral from the actual spiral did not significantly change during treatment **(Supplementary Figure 3)**. However, the movement appeared to be much smoother and consistent, with less staccato (starting and stopping) of the movement itself. As such, we quantified the difference in velocity during drawing and compared it to that of two age-matched healthy controls. Next, we quantified these movements before and after 6 months of NMN treatment. **Figure 6E** shows an example of one patient where the smoothness of her movement becomes more consistent after treatment. For four out of six patients, drawing movements at baseline were more jerky and less smooth compared to healthy controls **(Figure 6F).**

For these four patients, their smoothness improved significantly during treatment **(Figure 6G).** Two patients that remained stable in terms of smoothness after treatment, had already very smooth hand movements at baseline, and upon treatment stayed within the healthy control range.

Thus, instead of any clinical regression, which is part of the natural course of the disease^2^, patients experienced significant improvements in their movement disorder in terms of tremor, rigidity, ataxia and attention after 6 months of oral NMN supplementation at 250mg/day. These changes were reflected in the ICARS scores, which significantly dropped during the treatment period **(Figure 6H)**. Of note, three patients initially trialed higher doses of NMN (500, 750 or 1000 mg/day), however, none of these patients tolerated these doses due to increased tremor, hyperactivity and sleeping difficulties.

## Discussion

In this work, we studied cortical brain organoids of patients with pathogenic heterozygous *DHDDS* variants to assess pathophysiological mechanisms and novel treatment possibilities. We found that patient-derived cortical organoids showed a combination of developmental and degenerative abnormalities, characterized by mitochondrial dysfunction, a declining number of deep layer neurons, dolichol depletion, local cholesterol accumulation in astrocytes, glycosylation abnormalities and disorganized network activity. Next, we tested the potential of NAD+-enhancing therapy (Nicotinamide Mononucleotide, NMN), which was shown to rescue growth in a yeast model of DHDDS-related disease. Mitochondrial function and axon length could be restored upon Nicotinamide Mononucleotide treatment in cortical brain organoids. Similarly, six patients showed significant clinical improvements in an off-label N-of-1 trial with NMN. Together, these results reveal the underlying pathophysiology of heterozygous *DHDDS* variants and delineate a potential novel treatment option for this disease.

While glycosylation abnormalities have been found in patients with homozygous *DHDDS* variants^10^, patients with heterozygous *DHDDS* variants have normal transferrin profiles in blood.^9^ In mouse models with heterozygous *DHDDS* variants, glycosylation defects are evident both in circulating blood cells and in the brain.^11^ Here, we demonstrate that cortical brain organoids of human patients with heterozygous *DHDDS* variants do exhibit clear glycosylation abnormalities, challenging the assumption that these changes are restricted to homozygous disease. The glycosylation abnormalities in patients were progressive, which correlates with the progressive dolichol depletion that was observed in cortical brain organoids. An important next step will be to determine whether these glycosylation abnormalities improve following NMN treatment.

The specific localization of cholesterol in astrocytes is not surprising, as these cells are the primary producents of cholesterol in the brain.^16^ Astrocytes are strongly implicated in Niemann Pick C disease pathology^25^, and replenishing NPC1 solely in astrocytes alleviates the Niemann Pick C disease phenotypes in mice.^26^ Interestingly, NPC1 knockout in mice have increased dolichyl-P levels in liver homogenates, causing a compensatory decrease in the *de novo* synthesis of both dolichol and dolichyl-P.^27^ As such, DHDDS activity is severely reduced in these mice.^27^ These results suggest that the flux through the mevalonate pathway to cholesterol is linked to dolichol production and DHDDS activity directly. In addition, DHDDS and NPC2 were found to directly interact with each other^28^, and pathogenic *NUS1* variants lead to decreased NPC2 protein expression.^29^ However, in our cortical brain organoids, NPC2 expression was comparable between healthy controls and patients with *DHDDS* variants across the different timepoints. Together, these observations raise two non-mutually exclusive possibilities: cholesterol may influence DHDDS activity either through direct interactions with NPC proteins or indirectly via feedback on the mevalonate pathway.

Most patients with *DHDDS* variants exhibit a progressive disease course.^2^ While the pathophysiological mechanisms underlying this progression have remained elusive up until now, our work suggests that cortical organoids mimic this progression. They show morphological aberrancies starting around 4 months of age, which coincides with local cholesterol accumulation in astrocytes and gradual loss of deep-layer neurons. In addition, while dolichol and related molecules increase with time in healthy controls, they remain low in patients with *DHDDS* variants, and thus give rise to a (relative) progressive dolichol depletion. This concurs with progressive glycosylation abnormalities that were observed in patients. Intriguingly, the mitochondrial abnormalities observed were already present at earlier timepoints (day 90) and levels of developing neurons (ki67+) were decreased at early timepoints as well (day 60) in patients with *DHDDS* variants, suggesting that developmental issues also play a role in disease pathophysiology. Together, our results indicate that *DHDDS* variants lead to both developmental– and degenerative abnormalities.

Intriguingly, apart from NPC2, DHDDS was also found to interact with the mt-ND1 protein, part of mitochondrial complex I.^28^ Complex I dysfunction might explain our observation of mitochondrial dysfunction in fibroblasts and cortical brain organoids of patients with *DHDDS* variants. An alternative hypothesis for the mitochondrial dysfunction is cholesterol accumulation in mitochondria, which has also been observed in Niemann Pick C disease models and causes secondary mitochondrial dysfunction^30^. However, the mitochondrial dysfunction was already present in the organoids of patients with *DHDDS* variants before astrocytes – and cholesterol accumulation – developed. Regardless, we found that mitochondrial dysfunction worsens during later timepoints, and cholesterol accumulation could have contributed to that. Mitochondrial dysfunction at earlier time points could also be due to the fact that DHDDS variants causes glycosylation abnormalities. Glycosylation disorders such as PMM2-CDG, are similarly associated with secondary mitochondrial dysfunction.^31^

Multi-electrode array (MEA) revealed a more chaotic and immature bursting pattern in patients with *DHDDS* variants, as an increased number of spikes was produced outside actual burst. Nonetheless, the abnormalities on MEA were quite mild compared to other Congenital Disorders of Glycosylation (CDG).^13,32^ This might be due to the fact that other CDG have a clinical phenotype of severe intellectual disability and/or frequent seizures. In our DHDDS cohort, the patients were only mildly intellectually disabled and only patient one showed seizures during childhood, which resolved quickly under anti-epileptics. In mice, heterozygous *DHDDS* variants have been associated with decreased levels of interneurons.^11^ However, in organoids, interneurons only develop at much later timepoints than the timepoint assessed in this paper. Therefore, it might too early in organoid development to assess relevant pathophysiology. It would be interesting to see if the electrophysiological abnormalities worsen in patients around the time when interneurons normally develop.

In yeast harboring the DHDDS p.R205Q variant, several NAD+ precursors rescued growth. In human cortical brain organoids, treatment with NMN was able to completely reverse the mitochondrial phenotype of patients with heterozygous pathogenic *DHDDS* variants. A similar improvement of mitochondrial function has been observed in patient-derived cells of patients with mitochondrial diseases.^33^ This concurs with the beneficial clinical effects of niacin suppletion in mitochondrial myopathy patients.^34^ NMN or niacin leads to the inhibition of mTOR signaling and PPAR activation, which stimulates complex I activity and mitochondrial biogenesis. In addition, NMN is known to induce autophagy and lysosomal clearance.^35^ Thus, increased lysosomal clearance might explain the reduced lipid content, including cholesteryl esters, that was observed in CBOs of DHDDS patients after treatment. It would be interesting to assess whether NMN can reduce astrocyte-specific cholesterol content as well. However, if this is not the case, additional therapies might be needed to completely reverse the phenotype for DHDDS patients. This could involve the use of N-acetyl-leucine, which is beneficial for patients with Niemann Pick-C and helps lower cholesterol content.^36^ Similarly, miglustat can help lower cholesterol content in NPC patients and has been proven beneficial in fibroblasts of patients with heterozygous *DHDDS* variants.^12^

We assessed clinical improvements after NMN treatment using standardized outcome measures (ICARS) and newly developed outcome measures through the assessment of video material. While video assessments have not yet been used to assess therapeutic efficiency in CDG, it has been used to monitor treatment effects in Duchenne muscular dystrophy (DMD).^37^ AI-based assessment of walking and hand movement showed clear abnormalities in DMD patients in the pattern of movement trajectory, smoothness and symmetry of movement. In our cohort, we observed significant midline deviation during tandem walking and smoothness of hand movements in patients with *DHDDS* variants, which improved upon NMN treatment. The use of video-based assessments enables patients to record videos in a home setting and will allow patients to reduce travel time to clinical sites and doctor’s offices.

Together, we here found that heterozygous *DHDDS* variants give rise to a progressive, brain-specific disturbances that alter astrocyte lipid handling, mitochondrial function, and neuronal network stability in human cortical brain organoids. Although pathology was initially anticipated to be primarily neuronal, we identify a previously unrecognized astrocyte-restricted defect in cholesterol and phospholipid metabolism that emerges after gliogenesis and is accompanied by age-dependent vulnerability of deep-layer neurons. Integrated proteomic-, glycoproteomic-, and metabolomic analyses reveal progressive dolichol depletion and altered glycosylation of proteins involved in lysosomal and lipid trafficking, providing a mechanistic framework linking DHDDS dysfunction to impaired metabolic support of neuronal circuits. Modulation of cellular metabolism through NAD⁺ augmentation partially ameliorates lipid, mitochondrial and network phenotypes, supporting the concept that metabolic state influences disease expression and suggesting that DHDDS-related pathways may be relevant to neurodevelopmental and neurodegenerative phenotypes, including juvenile parkinsonism-plus, and merit further investigation in translational and preclinical contexts.

## Methods

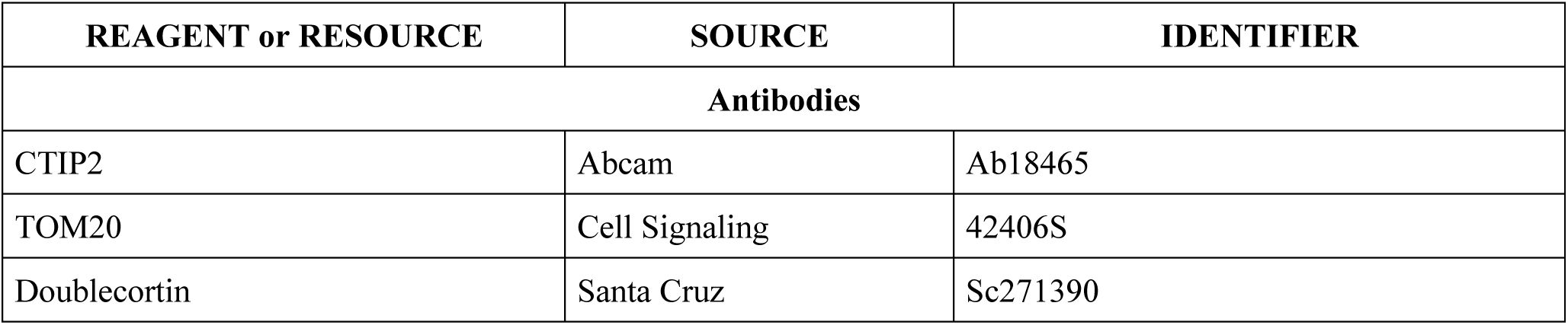
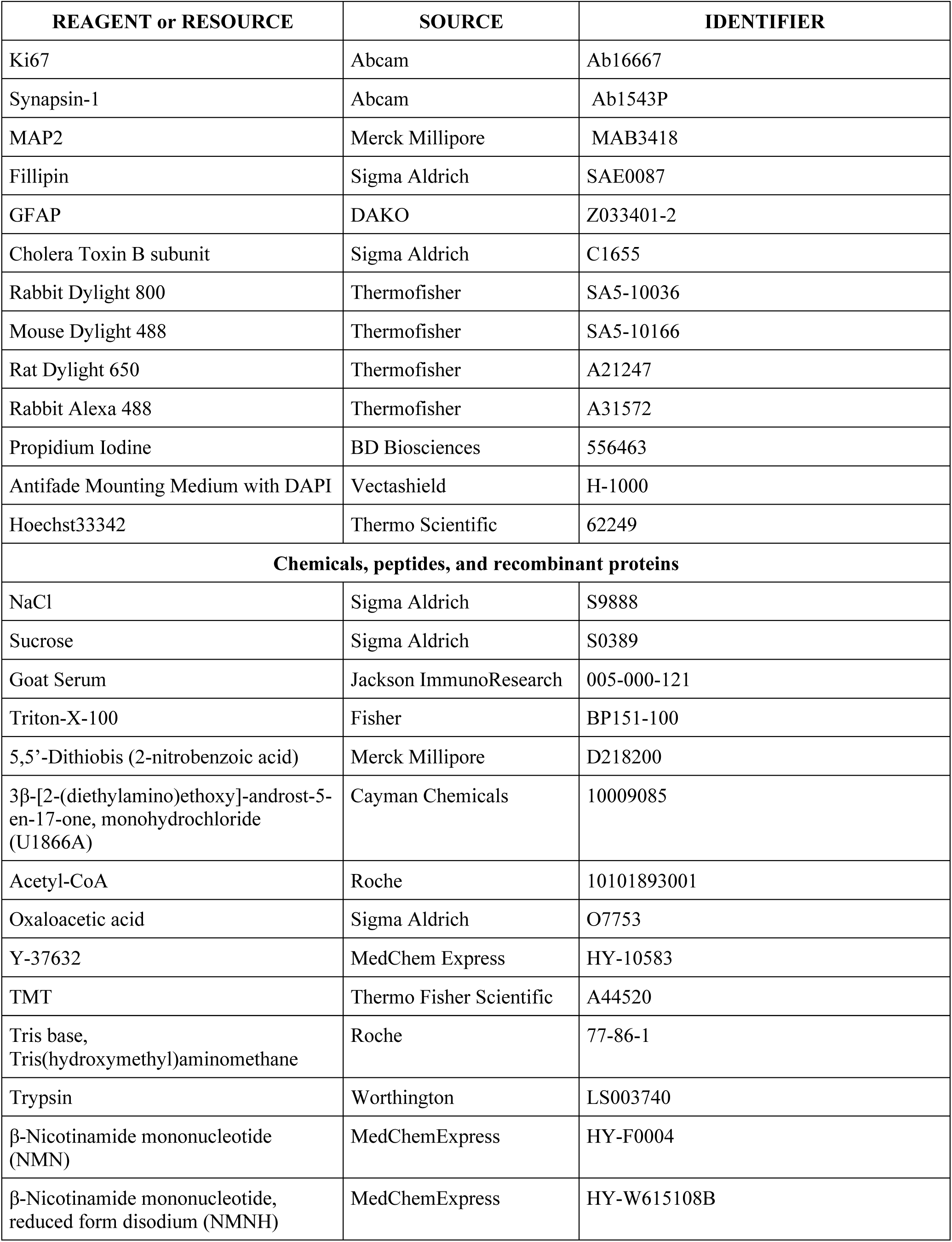
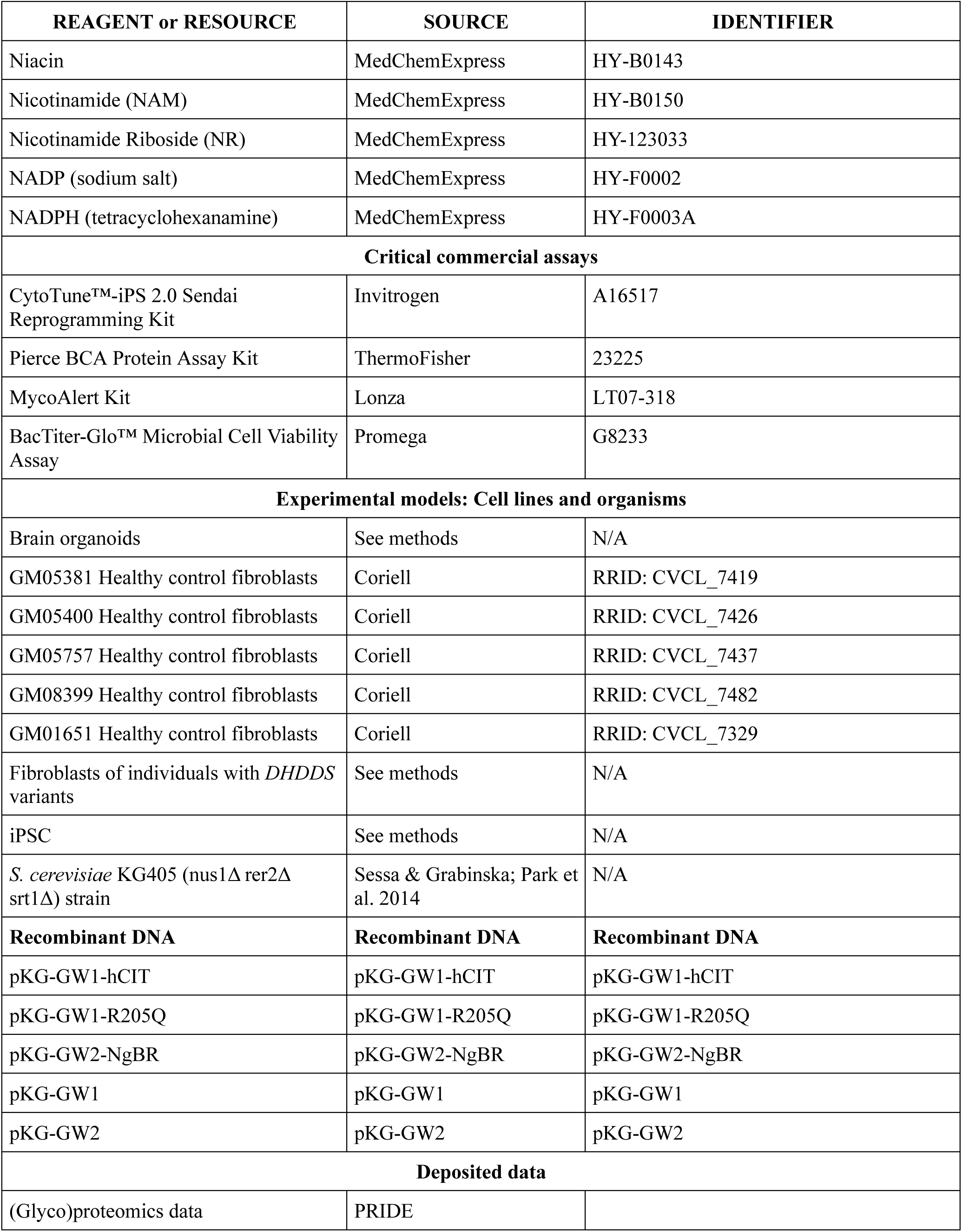
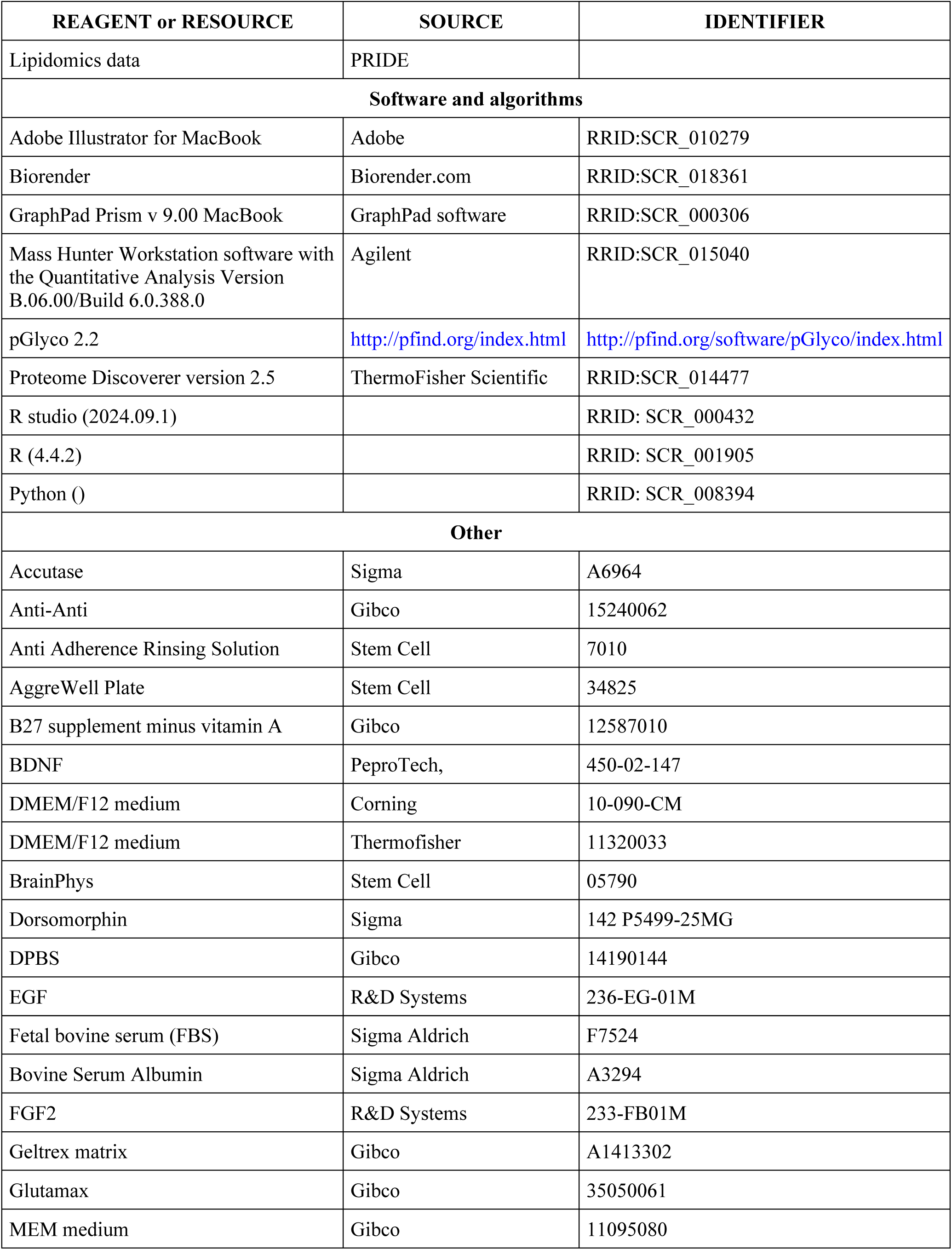
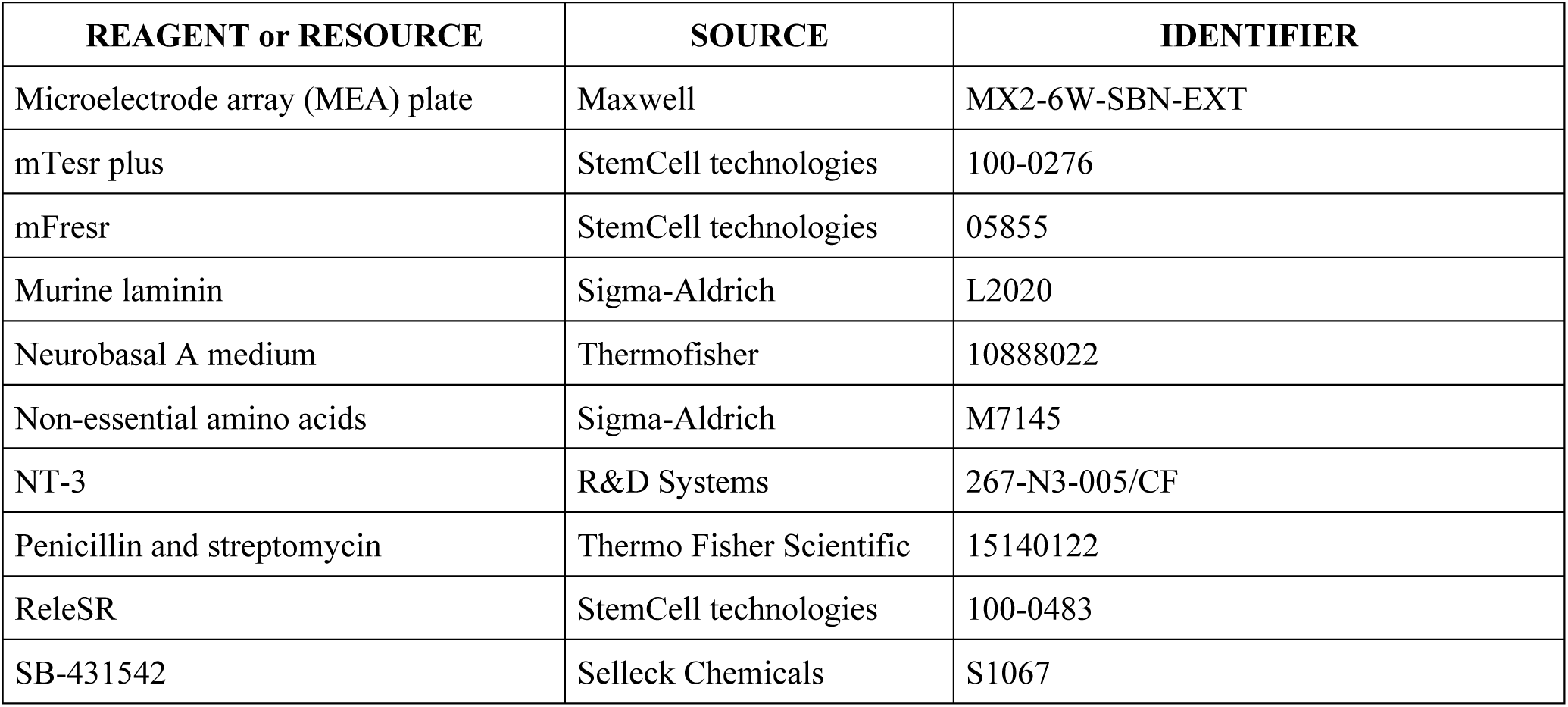

### Ethics

Informed research content was obtained from all patients included in the study. Samples from individuals with variants in *DHDDS* were collected during the Natural History Study (ClinicalTrials.gov Identifier: NCT04199000) in accordance with the Mount Sinai IRB (IRB: 23-00591). hiPSC cell lines were derived from fibroblasts collected from the patients (ethics application number S58358 and S60206 ‘‘Retrospective metabolomic analysis of archived fibroblasts’’ and Mayo Clinic IRB: 16–004682). Additional healthy fibroblasts were obtained from Coriell institute (GM05400, GM575, GM01651, GM05381, GM08399). Two hiPSC cell lines (1363, 8858) were a gift from Dr. Sergiu Pasca.

### Fibroblast Cultures

Fibroblasts from three patients were collected via skin-punch biopsy (Mayo IRB: 16–004682). Informed consent was obtained and recorded. Fibroblasts were maintained in MEM medium supplemented with 10% FBS and 1% Anti-Anti in the incubator at 37°C, 5% CO2. Routine mycoplasma testing was performed using the MycoAlert kit (Lonza).

### Generation of iPSCs

The fibroblast lines, as well as the age-matched controls, were reprogrammed into hiPSCs using the CytoTune™-iPS 2.0 Sendai Reprogramming Kit (Invitrogen A16517). Analysis of chromosomal abnormalities by karyotype G-banding was performed. Pluripotency of the derived hiPSC was assessed by flow cytometry, immunohistochemistry of the pluripotency markers (Oct 4, SSEA, Nanog, Tra-1-60), and three-germ layer (trilineage) differentiation (ectoderm, mesoderm, endoderm).

Mycoplasma testing was performed using MycoAlert kit (Lonza). hiPSC clones meeting all the criteria were selected and used for subsequent experiments. hiPSCs were maintained on reduced growth factor geltrex matrix coated plates (Gibco) in mTesr Plus medium (Stem Cell) supplemented with 10% mTeSR supplement (Stem Cell) and 1% Anti-Anti (Gibco) in the incubator at 37°C, 5% CO2.

### Generation of Cortical Organoids

Human cortical organoids were generated from iPSC.^39^ In short, once at 80–90% confluency, iPSC colonies were detached from plates to form spheroids using Accutase. The iPSCs were plated in an Aggrewell plate overnight, and the next day, they were placed in culture dishes. The spheroids were exposed to a series of small molecules to pattern toward dorsal forebrain identity. For the first 6 days in culture, spheroids were exposed to neural induction media (DMEM/F12, KSR, NEAA, Glutamax, Pen/Strep, Beta-mercaptoethanol) supplemented with Dorsomorphin (5 μM) and SB-431542 (10 μM) for dual-SMAD pathway inhibition. During this time, media was changed daily. Next, spheroids were treated with neural media, consisting of: neurobasal A, B-27 supplement without vitamin A, 1% GlutaMax, 1% penicillin and streptomycin, supplemented with EGF (20 ng/mL) and FGF2 (20 ng/mL). Neural media was changed daily through 15 days in culture, and then every other day for days 16–24. From days 25–43 in culture, spheroids were treated every other day with neural media supplemented with BDNF (20 ng/mL) and NT-3 (20 ng/mL). After day 43, maturing organoids were fed with neurobasal A only every 3–4 days until they were collected for experimental timepoints.

### Seahorse Respirometry

Fibroblasts were seeded 10,000 cells/well on 96 well XF Cell Culture Plates (Agilent) and cultured for 48 hours. Four wells were unseeded to establish baseline readings. For iPSCs, the plates were coated with geltrex and iPSCs were plated at 5,000 cells/well in mTesr+ medium with 10µM Y-27632 and grown until confluency. First, the plate was coated with poly-D-lysine. Next, the organoids were cut into 4 separate pieces, which were plated in spheroid microplates (Agilent). The seahorse assay was performed 1 hour after plating. Oxygen consumption and extracellular pH were assayed using the XFe96 Analyzer (Agilent) and the XF Cell Mito Stress Test kit (Agilent). Prior to the assay, standard culture media was replaced with XF Dulbecco’s Modified Eagle Media (DMEM) supplemented with 10 mM XF glucose, 10 mM XF pyruvate, and 10 mM XF glutamine. 2.5 μM mM oligomycin, 2 μM carbonylcyanide p-trifluoromethoxyphenylhydrazone (FCCP), a mixture of 500 nM rotenone and 500 nM antimycin A were administered sequentially to each well, and oxygen concentration and extracellular pH were measured three times following each administration. All OCR readouts were normalized to cell number and citrate synthase (CS) activity/abundance for the fibroblasts and iPSC experiments. For the organoids, OCR readouts were normalized against CS and protein concentration, measured using the Bicinchoninic Acid assay (BCA, Thermofisher).

Cell number was inferred by determining the number of nuclei per well. To determine nuclei number, wells were incubated in Hoechst 33342 nucleic acid stain (Thermo Scientific), and nuclei were counted using a Leice Widefield microscope at 10X. To determine CS concentration, cells were solubilized with 10 mM Tris-HCl + 0.3 % Triton X-100. To determine CS activity/abundance cell homogenates were incubated in a Tris-HCl-buffered solution in the presence of oxaloacetic acid, acetyl-CoA, and 5,5′-dithiobis-(2-nitrobenzoic acid) (DTNB). CoA-SH generated by the condensation of acetyl-CoA and oxaloacetic acid to citric acid by CS rapidly cleaves DTNB (colorless) to TNB−, which rapidly ionizes to TNB2− (yellow). Condensation of acetyl-CoA and oxaloacetic acid by CS was assayed via spectrophotometric measurement of TNB2− at 411 nm over 15 min.

### Immunohistochemistry

Organoids were fixed with 4% PFA for 1h10min at 4°C, washed with PBS, and then immersed in 30% sucrose for 24-48 hours. Organoids were embedded in O.C.T. after they sunk to the bottom of the tube containing 30% sucrose and O.C.T. blocks were frozen at −80C for cryosectioning. Sections of 10 µm thickness were washed in PBS, blocked for 15 min at room temperature (with 0.1% Triton X-100), and then incubated at 4°C overnight with primary antibodies diluted in blocking buffer solution. Primary antibodies used were CTIP2, MAP2, ki67, doublecortin, TOM20 and Synapsin-1. The next day, sections were washed with PBS and then incubated with secondary antibodies diluted in blocking buffer solution (2.5% donkey serum) for 1h at room temperature, protected from light. Nuclei were counterstained with Hoechts. Unbound secondary antibodies were washed with PBS and slides were coverslipped using Fluoromount-G mounting medium.

### Fillipin staining in fibroblasts, iPSCs and organoids

Fibroblasts were plated onto a LabTek chamber coverslide with 0.7cm^2^ surface per chamber at a density of 4,000 cells/well and cultured for 48 hours. iPSCs were plated on a geltrex-coated coverslide with 0.7cm^2^ surface at a density of 10,000 cells/well and grown to confluency for 48 hours. Organoids were sliced according to the “Immunohistochemistry” section and stained immediately. Slides were permeabilized with 0.1% triton-X-100 for 15 minutes, blocked with 1% BSA for 1 hour for iPSCs and fibroblasts, and with 4% BSA for 2 hours for organoids.

### Multi-electrode array

Electrophysiological recordings were conducted using the 6-well high-density Multi Electrode Array (MEA) MaxTwo system (HD-MEA) from MaxWell Biosystems. Wells were prepared following the MaxWell “Brain Organoid Plating Protocol, V2.0,” with minor modifications. Briefly, 6-well MEA plates were incubated in 1% Tergazyme solution for 2 hours at room temperature (RT), washed three times with distilled water, sterilized in ethanol for 30 minutes, and washed again three times with distilled water. Each well received 1 mL of Neurobasal-A (NBA) medium and was incubated for 48 hours for pre-conditioning.

After incubation, 50 µL of 0.07% poly(ethyleneimine) (PEI) was added to each well and incubated for 1 hour. Wells were then washed three times with distilled water and air-dried for 1 hour at RT. Next, 50 µL of 0.04 mg/mL laminin in NBA was added and incubated overnight. Laminin was aspirated, and cortical brain organoids were placed directly on the electrodes without media for 2–5 minutes. Subsequently, 50 µL of 0.04 mg/mL laminin in NBA was slowly added to each well, and the plates were incubated for 2 hours. For each organoid line, four MEA wells were prepared. After the 2-hour incubation, 1 mL of NBA was gently added to each well. The following day, half of the media was replaced with fresh NBA.

Organoids were maintained by changing the media every 3–4 days, and for the measurements, Neurobasal-A media was changed to BrainPhys media at least 24 hours before MEA recording. Recordings were performed using MaxLab software. The built-in “Activity Scan” protocol was used to identify active areas and assess firing rates. All parameters were kept at default except the recording duration, which was extended to 60 seconds to better detect slower activity. This was followed by the “Network Assay” protocol to evaluate neuronal network firing, using the neuronal units parameter. Lastly, the “Axon Tracking Assay” was performed using block parameters to detect axonal signal propagation.

Results were averaged across the six wells plated for each individual. The noise-normalized burst threshold was set at 2. Only wells with a firing rate >0.2 Hz were included for activity and network analysis.

### Proteomics and Glycoproteomics

Cortical organoids from individuals with *DHDDS* variants (N=3) and healthy controls (N=4) at Day 90 were collected and washed 3 times with DPBS, flash frozen in dry ice, and kept in −80 °C until the time of the assay. The samples were lysed using a Bioruptor sonication device in 8M urea (in 100 mM TEABC) with 1% protease inhibitor cocktail (Thermo Scientific). Protein amount was quantified in the organoid lysates using BCA colormetric assay as per the manufacturer’s instructions (Thermo Scientific). An equal quantity of protein was first reduced using 10 mM TCEP for 30 minutes at 55°C on a thermomixer, then alkylated with 40 mM iodoacetamide for 30 minutes in the dark at room temperature. The proteins were then digested, desalted, and tandem mass tags (TMT) labeled as previously described.^13^ Twenty percent of the peptides were fractionated using bRPLC for proteomics and 80% of the peptides were fractionated using size-exclusion chromatography (SEC) for glycoproteomics as previously described.^13^ LC-MS/MS for glycoproteomics and proteomics was conducted as previously described.^13^ Data analysis for glycoproteomics and proteomics was performed as described previously.^13^

### Lipidomics Analysis

Tissue homogenates were prepared in Milli-Q water using a Qiagen Tissuelyser II with 5 mm stainless steel beads (2×30 sec, 30 Hz). Protein concentration was measured via BCA assay.^40^ Lipidomics analysis, including lipid extraction, separation, and quantification, was performed as described by Vaz et al.^41^ In brief, lipids were extracted using 1:1 chloroform:methanol with internal standards for lipid major classes. Analysis was conducted on a Thermo Scientific Ultimate 3000 HPLC coupled to a Q Exactive Plus Orbitrap mass spectrometer, employing normal and reversed-phase chromatography in both negative and positive ion mode. Data processing and lipid annotation were performed using an in-house lipidomics pipeline in R and MATLAB, as previously described.^41^ Lipid identification was based on a combination of accurate mass, predicted retention times, previous analyses of samples with known metabolic defects, and the injection of relevant standards. Specifically, for dolichol-type lipids (dolichol, polyprenol, polyprenal, dolichoic acid and dolichol esters) a cubic B-spline function was used to predict the retention time as a function of the dolichol chain length. Predictions were similar for the different dolichol types, showing the same relation between retention time and dolichol chain length but with different intercept. The presence of the identified dolichol lipid clusters at the predicted retention times was confirmed in several in-house mass spectrometry datasets.

### Assessment of clinical parameters before and after NMN treatment

Patients were asked to record home videos before starting NMN treatment and after 4-6 months of 250mg NMN treatment. Videos included tandem walking, for which parents taped a line to the ground which patients had to walk on. Patients were instructed to move arms and legs as minimally as possible and stay close to line, while walking in tandem position. The videos were analyzed in ImageJ using the Manual Tracking Plug In tool, which was placed at the top of the head, the midline, both arms and feet. The midline was also tracked during the tandem walk, and x-values of the line were subtracted from the x-values of the arms and feet. The deviation in pixels was normalized against the patient height, which was captured in each frame. This normalization ensured that apparent differences in movement amplitude were not due to patients walking closer to– or farther away from the camera but reflected true movement deviations.

### Yeast strains and plasmids

All *Saccharomyces cerevisiae* strains were derived from the S288C (BY4742) background. Humanized yeast strains were generated using the triple-deletion strain KG405 (nus1Δ rer2Δ srt1Δ)^42^, which carries a URA3-marked plasmid expressing *Giardia lamblia* cis-prenyltransferase (pGlcis-PTase-URA) for viability. Yeast strains and plasmids were kindly provided by Dr. William Sessa and Dr. Kariona Grabinska.

KG405 cells were co-transformed with pKG-GW1 vectors expressing human DHDDS (wild-type: pKG-GW1-hCIT or p.R205Q variant: pKG-GW1-R205Q) and pKG-GW2 vectors expressing wild-type human NgBR (pKG-GW2-NgBR), as previously described.^9^ DHDDS constructs were selected using LEU2, and NgBR was selected using MET17. Control strains expressed wild-type DHDDS and NgBR (WT) or empty pKG-GW1 and pKG-GW2 vectors (vector control). To render growth dependent on human cis-prenyltransferase activity, transformants were counterselected on 5-fluoroorotic acid (FOA; *1 g L⁻¹*) to eliminate the URA3-marked pGlcis-PTase plasmid. Transformants were maintained in synthetic dropout medium lacking uracil, leucine and methionine at 30°C.

Luminescence-based viability measurements for growth assays, screening and dose-response validation were performed using the BacTiter-Glo Microbial Cell Viability Assay.

### High-throughput compound screening

High-throughput screening was performed using the L4000 Bioactive Compound Library (TargetMol; 8,387 compounds). Compounds or DMSO vehicle were dispensed into 384-well plates at 20 µM final concentration (0.72% DMSO) using an Echo 650 acoustic dispenser (Beckman Coulter). Overnight cultures of DHDDS p.R205Q-expressing and wild-type complemented yeast were diluted in SD–Leu–Met medium containing 5-fluoroorotic acid (FOA) to an OD600 of 0.0025 and dispensed into plates using an EL406 automated dispenser. Plates were incubated at 30°C for 24 h, after which ATP-based luminescence was measured by adding BacTiter-Glo reagent at a 1:1 volume ratio. Plates were incubated for 20 min and read using an EnVision plate reader (PerkinElmer). Screening was performed in singlicate, and each plate included multiple wells of DHDDS p.R205Q mutant cells (negative control) and wild-type complemented cells (positive control).

### Yeast dose-response assays

For dose–response validation, DHDDS p.R205Q-expressing yeast were treated with increasing concentrations of NADP, NADPH, or NAD⁺-related compounds (niacin, NMN, NMNH, NR, and NAM) in 384 well plates (0.72% DMSO final). After incubation at 30°C for 24 h, ATP-based luminescence was measured using BacTiter-Glo reagent. Each concentration was tested in triplicate, and DMSO vehicle controls were assayed in 16 technical replicates.

### Spiral Analysis

Hand-drawn spiral images (JPG/PNG) and PDFs (first page only) were quantified to assess how closely each hand-drawn tracing followed the printed Archimedes spiral. Analyses were implemented in Python using NumPy (version 2.3, array operations, FFT), OpenCV (Version 4.10.x, image I/O, color conversion, resizing), scikit-image (version 0.24, Sobel edge detection for fallback segmentation), PyMuPDF/fitz (version 1.26, PDF rasterization), and pandas (version 2.2, tabulation/CSV export). For consistency across scanners and cameras, large images were temporarily resized to ∼800 px on the long edge; all geometric measures were then scaled back to original units. The printed reference spiral (gray/black) and the participant’s drawing (typically colored) were separated using color thresholds, with an edge-based fallback for dark pens. Table Sx summarizes each tracing’s deviation from the ideal spiral—reported as mean absolute deviation (MAD), standard deviation (SD), and maximum deviation (MaxD)—and the data volume used to compute these metrics, including n_ref (pixels from the printed/reference spiral after masking/subsampling) and n_child (pixels from the participant’s drawing).

### Data Analysis and Statistical Methods

Graphs were created with GraphPad Prism, R and R-studio. Images were analyzed with ImageJ. Gene Set Enrichment Analysis (GSEA) was performed using the ClusterProfiler package in R (version 4.12.6) using the GO Biological Process Ontology or the MitoCarta database.

## Data Availability

All data produced in the present work are contained in the manuscript

## Acknowledgements

We thank Arie Dane for the bioinformatic analysis of metabolomics data. We thank the Advanced Bioimaging Core at Mount Sinai for the use of their microscopes. We thank the UC Berkeley QB3 High-Throughput Screening Facility and the UCSF Chemical Screening Center for technical support with high-throughput screening and compound library preparation. This study was supported by 1U54NS115198-01 from the National Institute of Neurological Diseases and Stroke (NINDS), the National Center for Advancing Translational Sciences (NCATS), the National Institute of Child Health and Human Development (NICHD), and the Rare Disorders Consortium Disease Network (RDCRN).

