## Supplementary Figure 1 for "DHDDS-related juvenile parkinsonism is caused by impaired lipid metabolism, glycosylation, and mitochondrial dysfunction, which can be rescued by NAD⁺ treatment"

**Supplementary Files**

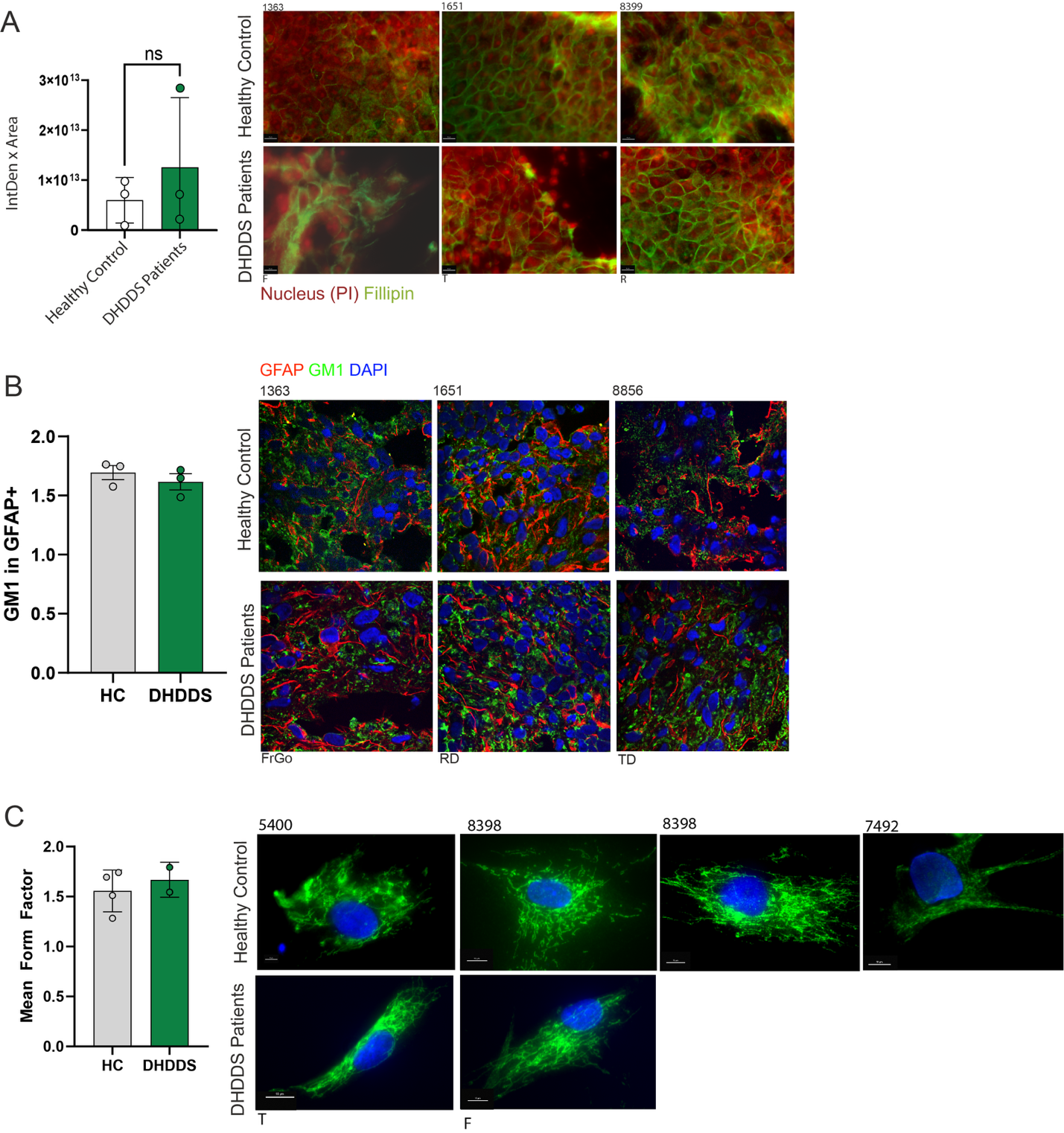

**Supplementary Figure 1:**

A. Fillipin staining in iPSCs of patients with *DHDDS* variants with and without U-1866A treatment. Fillipin intensity was quantified by calculating the integrated density (IntDen) values with ImageJ and multiplying this by the Fillipin total area. Healthy controls (N=4), patients with *DHDDS* variants (N=3). For each individual, at least three different images were quantified.
B. GM1 staining in astrocytes (GFAP) and whole organoid sections. The bars reflect the mean ratio of GM1 in astrocytes / GM1 intensity in the entire organoid ±SEM. Students t-test was used to calculate significance. p>0.05.
C. TOM20 staining in fibroblasts of patients with *DHDDS* variants (N=2) and healthy controls (N=4). Form factor was calculated with ImageJ plugin Mitochondria-Analyzer.^40^ The mean form factor by averging the form factor of 4 images per individual. The bar graphs show the mean form factor for all individuals for the two groups ±SD.

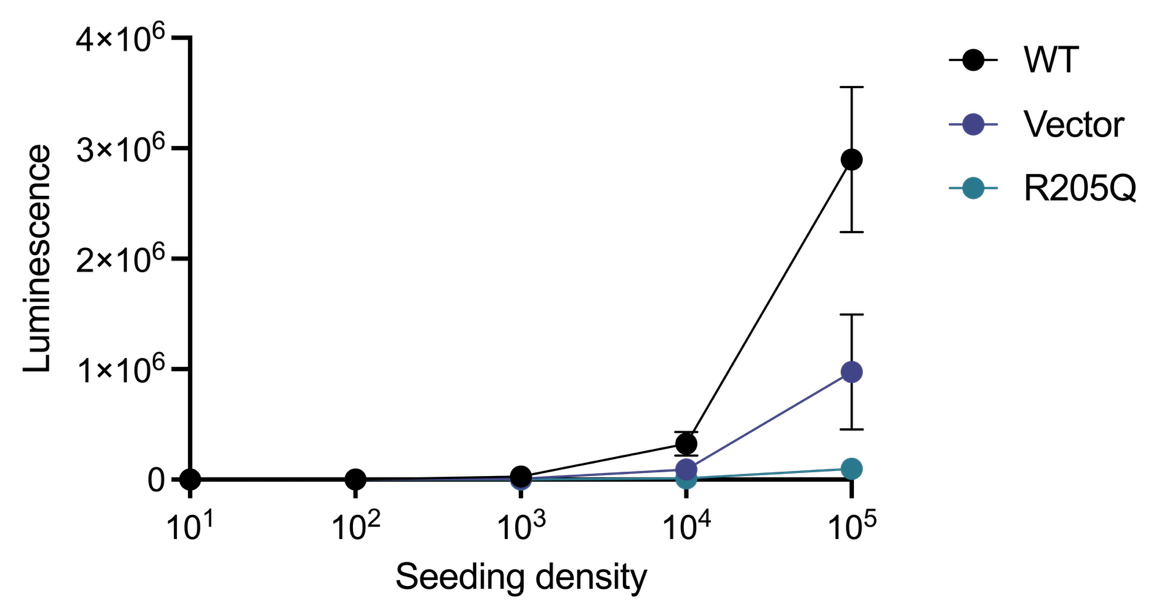

**Supplementary Figure 2:** Growth analysis of humanized yeast strains expressing wild-type and p.R205Q DHDDS. *Rer2/ srt1 nus1* knockout yeast yeast were complemented with wild-type DHDDS and NgBR (WT), empty vectors (vector), or p.R205Q DHDDS together with wild-type NgBR (R205Q). Growth was assessed by ATP-based luminescence following serial dilution and 24 h incubation. Data represent mean ± SD (n=3 technical replicates).

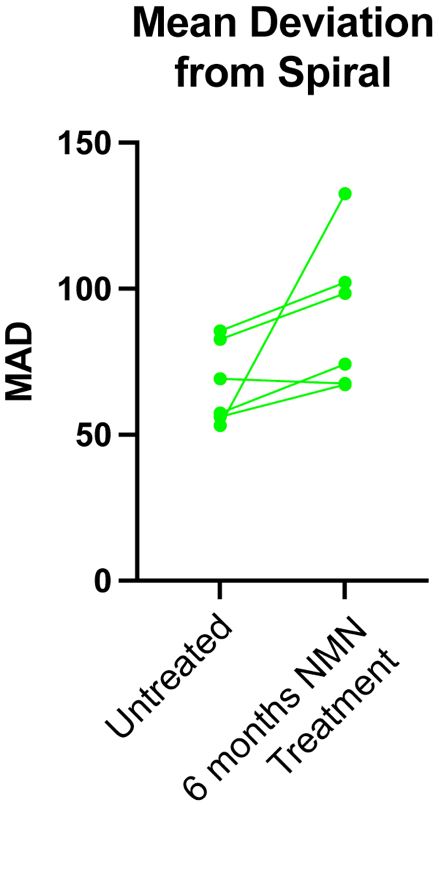

**Supplementary Figure 3:** Showing the mean deviation (MAD) of the patient-drawn Archimedes spiral from the reference spiral in patients before NMN treatment and during NMN treatment.
